# Assessing the Impact of the Healthy Vaccinee Effect on COVID-19 Vaccine Effectiveness Estimates

**DOI:** 10.64898/2026.01.02.26343330

**Authors:** Hiam Chemaitelly, Houssein H. Ayoub, Peter Coyle, Patrick Tang, Mohammad R. Hasan, Hadi M. Yassine, Asmaa A. Al Thani, Zaina Al-Kanaani, Einas Al-Kuwari, Andrew Jeremijenko, Anvar Hassan Kaleeckal, Ali Nizar Latif, Riyazuddin Mohammad Shaik, Hanan F. Abdul-Rahim, Gheyath K. Nasrallah, Mohamed Ghaith Al-Kuwari, Hamad Eid Al-Romaihi, Mohamed H. Al-Thani, Abdullatif Al-Khal, Roberto Bertollini, Adeel A. Butt, Laith J. Abu-Raddad

## Abstract

**Background:** Health status influences uptake of COVID-19 vaccination, with vaccinated individuals generally being healthier than their unvaccinated counterparts. However, the extent to which this healthy vaccinee effect biases vaccine effectiveness estimates remains unclear. This study investigated how different levels of the healthy vaccinee effect may impact these estimates.

**Methods:** Three national, matched, retrospective cohort studies were conducted on the population of Qatar from February 5, 2020-May 14, 2024, to estimate effectiveness of primary-series and booster vaccinations and of natural infection against infection and reinfection. The studies explored three different scenarios for matching by coexisting conditions, each reflecting a different level of health-status imbalance between vaccinated and unvaccinated individuals.

**Results:** Estimated effectiveness of primary-series and booster vaccinations against infection and against severe, critical, or fatal COVID-19 showed consistent values across the different matching strategies, despite varying hazard ratios for non-COVID-19 deaths. For example, primary-series effectiveness against severe, critical, or fatal COVID-19 was 96.3% (95% CI: 94.5-97.5) when matching by exact coexisting condition types, 94.2% (95% CI: 92.5-95.6) when matching by number of coexisting conditions, and 94.0% (95% CI: 92.3-95.3) with no matching by coexisting conditions. Similar results were observed when the analysis was stratified by the duration of follow-up and for the subgroup of individuals clinically vulnerable to severe COVID-19. Analogous results were found for effectiveness of natural infection against reinfection and against severe, critical, or fatal COVID-19 upon reinfection.

**Conclusion:** The imbalance in health status between vaccinated and unvaccinated individuals in real-world conditions did not introduce a healthy vaccinee bias into the estimated vaccine effectiveness.

## Introduction

Health status influences the uptake of preventive health measures, particularly among the elderly [1, 2]. Healthier elderly individuals are more likely to seek vaccinations, whereas those with poorer health face barriers such as limited mobility, severe illnesses, and frailty [1, 2]. This disparity in vaccination uptake can result in the "healthy vaccinee effect" [1, 3].

The healthy vaccinee effect occurs when healthier individuals, who naturally experience better health outcomes, are more likely to be vaccinated, potentially leading to an overestimation of vaccine effectiveness [1, 3]. This introduces what is known as the "healthy vaccinee bias" [1, 3]. The healthy vaccinee effect has been documented in influenza [1, 3–5] and coronavirus disease 2019 (COVID-19) vaccine effectiveness studies [6–9], where strong protective effects against death not attributed to influenza or COVID-19 have been observed, despite these vaccines not being expected to influence such outcomes.

In a recent national study on Qatar’s population, we identified strong evidence for the healthy vaccinee effect, demonstrating substantial protection from vaccination against non-COVID-19 death [9]. This effect was observed only in the first six months post-vaccination and among individuals aged over 50 years and those more clinically vulnerable to severe COVID-19 [9]. The effect was attributed to higher short-term mortality among seriously ill, frail, and less mobile individuals, who are less likely to receive the vaccine [9].

Building on this research, this national retrospective cohort study on the same population investigated whether the documented healthy vaccinee effect can bias the estimation of vaccine effectiveness of primary-series and booster vaccinations against SARS-CoV-2 infection and against severe, critical, or fatal COVID-19. To assess this, vaccine effectiveness was estimated and compared across three cohort matching strategies based on coexisting conditions: exact matching by condition types, matching by the number of conditions, and no matching. These strategies represent varying levels of the healthy vaccinee effect, i.e., different levels of health-status imbalance between vaccinated and unvaccinated individuals. The analyses were further extended to evaluate the impact of these matching strategies on estimating the effectiveness of natural infection in protecting against reinfection and against severe, critical, or fatal COVID-19 upon reinfection.

## Materials and methods

### Study population, data sources, and vaccination

This study encompassed Qatar’s resident population from February 5, 2020, the date of the earliest SARS-CoV-2 test, to May 14, 2024, the study’s end date. Data on COVID-19 laboratory testing, vaccination, hospitalization, and death were sourced from Qatar’s national integrated digital health information platform (Supplementary Section 1 in Supplementary Appendix).

The national platform includes all SARS-CoV-2-related data, including COVID-19 vaccinations, hospitalizations, and polymerase chain reaction (PCR) and medically supervised rapid antigen tests, irrespective of location or facility (Supplementary Section 2). Testing in Qatar was extensive, with most infections identified through routine testing rather than symptomatic presentation (Supplementary Section 1) [10–12]. The national platform also includes data on coexisting conditions for individuals based on the health record encounters of each individual in the national public healthcare system (Supplementary Section 3).

COVID-19 vaccinations in Qatar were administered primarily using mRNA vaccines [12–14], and were provided free of charge to all individuals, irrespective of citizenship status, exclusively through the public healthcare system [15]. The vaccination strategy prioritized frontline healthcare workers, individuals with severe or multiple chronic conditions, and individuals aged over 50 years [10].

Demographic information was obtained from the national health registry. Qatar’s demographic structure is distinct. Only 9% of the population are over 50 years and 89% are resident expatriates from over 150 countries [16]. Further details on Qatar’s population and COVID-19 databases have been previously published [10, 11, 16–20].

### Study design

Three national, matched, retrospective cohort studies were conducted, informed by our earlier studies on Qatar’s population [12, 17, 19, 21–25]. Each study compared effectiveness estimates across three scenarios for matching by coexisting conditions: exact matching by coexisting condition types, matching by number of coexisting conditions, and no matching by coexisting conditions.

The first study (two-dose analysis) compared the incidence of infection and severe, critical, or fatal COVID-19 in the national cohort of individuals who received the primary-series vaccination (two-dose cohort) with those in the national cohort of unvaccinated individuals (unvaccinated cohort). The second study (three-dose analysis) compared these outcomes in the national cohort of individuals who received a third (booster) vaccine dose (three-dose cohort) with those in the two-dose cohort. The third study (natural-infection analysis) compared the incidence of reinfection and severe, critical, or fatal COVID-19 upon reinfection in the national cohort of individuals with a documented primary infection (primary-infection cohort) with those in the national cohort of uninfected individuals (uninfected cohort). All studies assessed the incidence of non-COVID-19 deaths across study cohorts to measure the healthy vaccinee effect [9].

Incidence of infection was defined as a SARS-CoV-2-positive test after the start of follow-up. Individuals whose infection progressed to severe, critical, or fatal COVID-19 were classified based on their worst outcome, starting with COVID-19 death [26], followed by critical disease [27], and then severe disease [27] (Supplementary Section 4). Infection severity was determined by trained medical personnel independent of study investigators. Classifications were based on individual chart reviews, following the World Health Organization (WHO) guidelines for defining COVID-19 severity (acute-care hospitalization) [27], criticality (intensive-care-unit hospitalization) [27], and fatality [26] (Supplementary Section 4) [28].

Non-COVID-19 deaths were sourced from the national federated mortality database, which captures all deaths in healthcare facilities and elsewhere, including forensic deaths investigated by Qatar’s Ministry of Interior [20]. All deaths not classified as COVID-19 deaths were considered non-COVID-19 deaths.

### Inclusion criteria and matching

Inclusion in the vaccine effectiveness studies required having two doses of an mRNA vaccine for the two-dose cohort, and three doses for the three-dose cohort. Those who received the ChAdOx1 nCoV-19 (AZD1222) vaccine, a small part of the population, or the pediatric 10-µg BNT162b2 vaccine [22] were excluded. The unvaccinated cohort included individuals with no vaccination record at the start of follow-up.

Inclusion in the natural-infection effectiveness study required having a record of primary infection for the primary-infection cohort. Individuals who received the pediatric 10-µg BNT162b2 vaccine [22] or a mix of different vaccine types before the start of follow-up were excluded.

Cohorts were matched exactly one-to-one by sex, 10-year age group, and nationality. Vaccine effectiveness studies also included matching by prior documented SARS-CoV-2 infection status (no prior infection, prior pre-omicron infection, prior omicron infection, or prior pre-omicron and omicron infections). Analogously, the natural-infection effectiveness study included matching by prior vaccine type and number of vaccine doses. Matching by calendar time was further implemented to ensure that matched pairs were concurrently present in Qatar (Supplementary Section 5). Matching by these factors aimed to balance confounders across cohorts, as history of infection or vaccination may affect an individual’s health status [29], susceptibility to infection [30], or vaccination uptake [31].

Iterative matching was implemented in vaccine effectiveness studies to ensure that, at the start of follow-up, individuals were alive, had maintained their vaccination status, had the same prior infection status as their match, and had no documented SARS-CoV-2 infection within the previous 90 days. The 90-day duration was used to avoid misclassifying a previous prolonged SARS-CoV-2 infection as an incident infection [32–34]. In the natural-infection effectiveness study, iterative matching ensured that, at the start of follow-up, individuals were alive, had maintained their prior infection status, and had the same vaccine type and number of doses as their match. Further details can be found in Supplementary Section 5.

### Cohorts’ follow-up

Follow-up started from the date of the second vaccine dose in the two-dose analysis, date of the third vaccine dose in the three-dose analysis, and 90 days after the primary infection in the natural-infection analysis. For exchangeability [17, 35], both members of each matched pair were censored at the earliest occurrence of receiving an additional vaccine dose.

Accordingly, individuals were followed until the first of any of the following events: a documented SARS-CoV-2 infection (irrespective of symptoms), a new vaccine dose (with matched-pair censoring), death, or the administrative end of follow-up at the end of the study.

### Oversight

The institutional review boards at Hamad Medical Corporation and Weill Cornell Medicine– Qatar approved this retrospective study with a waiver of informed consent. The study was reported according to the Strengthening the Reporting of Observational Studies in Epidemiology (STROBE; Supplementary Table 1).

### Statistical analysis

Matched cohorts were characterized through descriptive statistics and compared using standardized mean differences (SMDs). An SMD of ≤0.1 indicated adequate matching[36].

Incidence rates of each of infection, severe, critical, or fatal COVID-19, and non-COVID-19 death, defined as number of individuals with the outcome of interest in a cohort divided by number of person-weeks contributed by all individuals in the cohort, were estimated with their associated 95% confidence intervals (CIs), using Poisson log-likelihood regression models.

Overall adjusted hazard ratios (aHRs) comparing the incidence of each outcome of interest between study cohorts, and corresponding 95% CIs, were calculated using Cox regression models with adjustment for the matching factors. This adjustment ensured unbiased estimation of the standard variance [37]. The aHRs comparing the incidence of SARS-CoV-2 infection and severe, critical, or fatal COVID-19 between cohorts were further adjusted for SARS-CoV-2 testing rates. To explore differences in the risk of severe, critical, or fatal COVID-19 over time, the aHRs were estimated before and after 6 months from the start of follow-up using separate Cox regressions, with "failure" restricted to specific time intervals. An additional analysis was conducted, including only individuals clinically vulnerable to severe COVID-19, defined as those aged over 50 years or with two or more coexisting conditions [19]. CIs were not adjusted for multiplicity. Interactions were not investigated.

Effectiveness estimates (and 95% CIs) were calculated as 1-aHR if the aHR was <1, and as 1/aHR-1 if the aHR was ≥ 1 [19, 38]. This approach ensured a symmetric scale for both negative and positive effectiveness, spanning from -100%-100%, resulting in a meaningful interpretation of estimates [19, 38].

Statistical analyses were performed using Stata/SE version 18.0 (Stata Corporation, College Station, TX, USA).

## Results

### Two-dose analysis

Supplementary Fig. 1 illustrates the matched cohorts’ selection process. Table 1 describes the cohorts’ baseline characteristics. Cohorts comprised 811,944 individuals matched by coexisting condition types, 845,469 matched by the number of coexisting conditions, and 863,488 not matched by coexisting conditions.

**Table 1.**
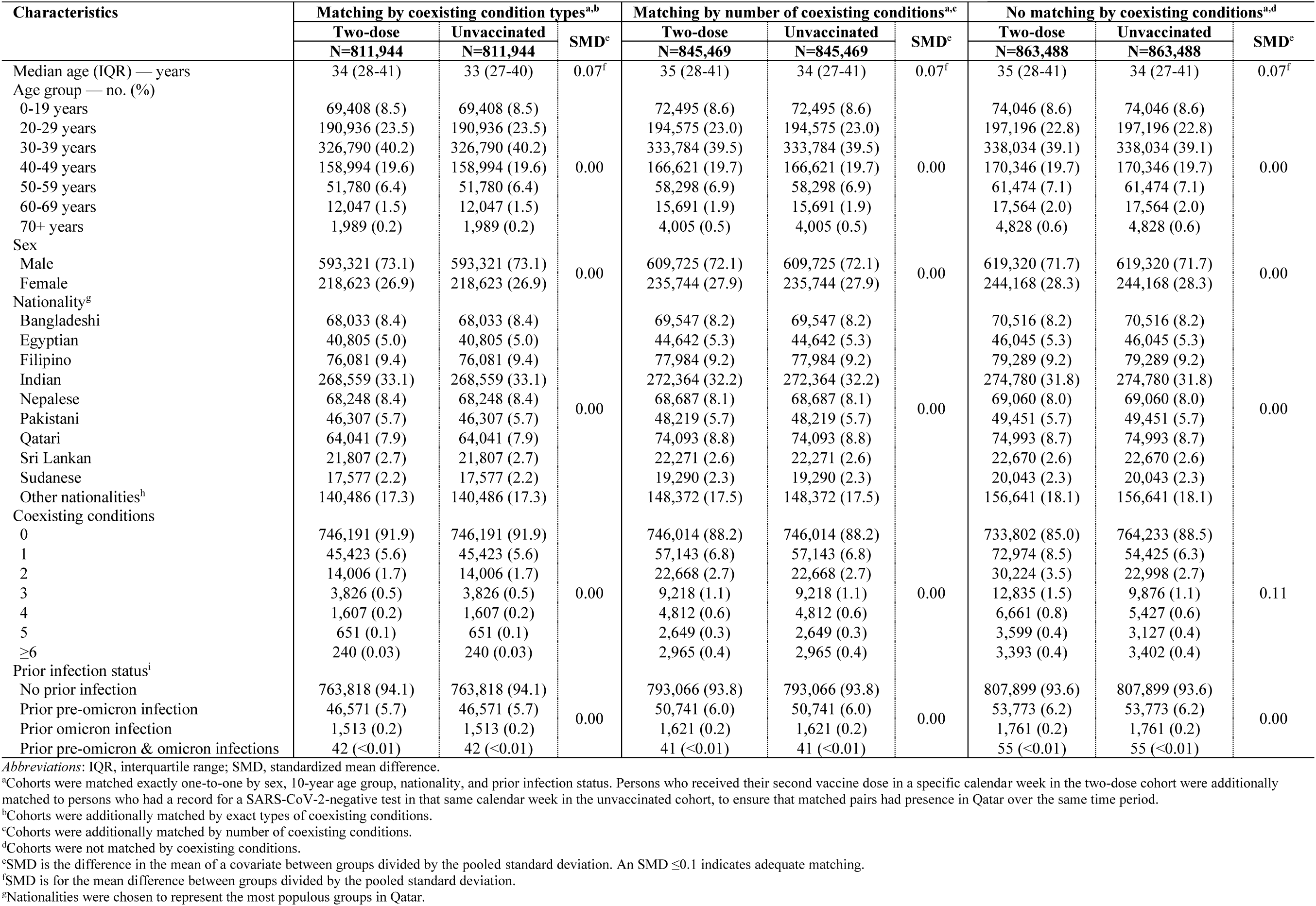
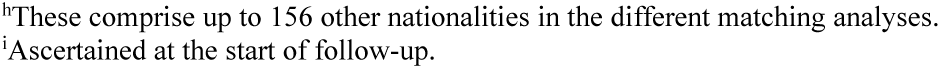
Baseline characteristics of matched cohorts in the two-dose (primary-series) analysis.

| Characteristics | Matching by coexisting condition types<sup>a,b</sup> |  |  | Matching by number of coexisting conditions<sup>a,c</sup> |  |  | No matching by coexisting conditions<sup>a,d</sup> |  |  |
| --- | --- | --- | --- | --- | --- | --- | --- | --- | --- |
|  | Two-dose | Unvaccinated | SMD<sup>e</sup> | Two-dose | Unvaccinated | SMD<sup>e</sup> | Two-dose | Unvaccinated | SMD<sup>e</sup> |
|  | N=811,944 | N=811,944 |  | N=845,469 | N=845,469 |  | N=863,488 | N=863,488 |  |
| Median age (IQR) — years | 34 (28-41) | 33 (27-40) | 0.07<sup>f</sup> | 35 (28-41) | 34 (27-41) | 0.07<sup>f</sup> | 35 (28-41) | 34 (27-41) | 0.07<sup>f</sup> |
| Age group — no. (%) |  |  |  |  |  |  |  |  |  |
| 0-19 years | 69,408 (8.5) | 69,408 (8.5) |  | 72,495 (8.6) | 72,495 (8.6) |  | 74,046 (8.6) | 74,046 (8.6) |  |
| 20-29 years | 190,936 (23.5) | 190,936 (23.5) |  | 194,575 (23.0) | 194,575 (23.0) |  | 197,196 (22.8) | 197,196 (22.8) |  |
| 30-39 years | 326,790 (40.2) | 326,790 (40.2) |  | 333,784 (39.5) | 333,784 (39.5) |  | 338,034 (39.1) | 338,034 (39.1) |  |
| 40-49 years | 158,994 (19.6) | 158,994 (19.6) | 0.00 | 166,621 (19.7) | 166,621 (19.7) | 0.00 | 170,346 (19.7) | 170,346 (19.7) | 0.00 |
| 50-59 years | 51,780 (6.4) | 51,780 (6.4) |  | 58,298 (6.9) | 58,298 (6.9) |  | 61,474 (7.1) | 61,474 (7.1) |  |
| 60-69 years | 12,047 (1.5) | 12,047 (1.5) |  | 15,691 (1.9) | 15,691 (1.9) |  | 17,564 (2.0) | 17,564 (2.0) |  |
| 70+ years | 1,989 (0.2) | 1,989 (0.2) |  | 4,005 (0.5) | 4,005 (0.5) |  | 4,828 (0.6) | 4,828 (0.6) |  |
| Sex |  |  |  |  |  |  |  |  |  |
| Male | 593,321 (73.1) | 593,321 (73.1) | 0.00 | 609,725 (72.1) | 609,725 (72.1) | 0.00 | 619,320 (71.7) | 619,320 (71.7) | 0.00 |
| Female | 218,623 (26.9) | 218,623 (26.9) |  | 235,744 (27.9) | 235,744 (27.9) |  | 244,168 (28.3) | 244,168 (28.3) |  |
| Nationality<sup>g</sup> |  |  |  |  |  |  |  |  |  |
| Bangladeshi | 68,033 (8.4) | 68,033 (8.4) |  | 69,547 (8.2) | 69,547 (8.2) |  | 70,516 (8.2) | 70,516 (8.2) |  |
| Egyptian | 40,805 (5.0) | 40,805 (5.0) |  | 44,642 (5.3) | 44,642 (5.3) |  | 46,045 (5.3) | 46,045 (5.3) |  |
| Filipino | 76,081 (9.4) | 76,081 (9.4) |  | 77,984 (9.2) | 77,984 (9.2) |  | 79,289 (9.2) | 79,289 (9.2) |  |
| Indian | 268,559 (33.1) | 268,559 (33.1) |  | 272,364 (32.2) | 272,364 (32.2) |  | 274,780 (31.8) | 274,780 (31.8) |  |
| Nepalese | 68,248 (8.4) | 68,248 (8.4) | 0.00 | 68,687 (8.1) | 68,687 (8.1) | 0.00 | 69,060 (8.0) | 69,060 (8.0) | 0.00 |
| Pakistani | 46,307 (5.7) | 46,307 (5.7) |  | 48,219 (5.7) | 48,219 (5.7) |  | 49,451 (5.7) | 49,451 (5.7) |  |
| Qatari | 64,041 (7.9) | 64,041 (7.9) |  | 74,093 (8.8) | 74,093 (8.8) |  | 74,993 (8.7) | 74,993 (8.7) |  |
| Sri Lankan | 21,807 (2.7) | 21,807 (2.7) |  | 22,271 (2.6) | 22,271 (2.6) |  | 22,670 (2.6) | 22,670 (2.6) |  |
| Sudanese | 17,577 (2.2) | 17,577 (2.2) |  | 19,290 (2.3) | 19,290 (2.3) |  | 20,043 (2.3) | 20,043 (2.3) |  |
| Other nationalities<sup>h</sup> | 140,486 (17.3) | 140,486 (17.3) |  | 148,372 (17.5) | 148,372 (17.5) |  | 156,641 (18.1) | 156,641 (18.1) |  |
| Coexisting conditions |  |  |  |  |  |  |  |  |  |
| 0 | 746,191 (91.9) | 746,191 (91.9) |  | 746,014 (88.2) | 746,014 (88.2) |  | 733,802 (85.0) | 764,233 (88.5) |  |
| 1 | 45,423 (5.6) | 45,423 (5.6) |  | 57,143 (6.8) | 57,143 (6.8) |  | 72,974 (8.5) | 54,425 (6.3) |  |
| 2 | 14,006 (1.7) | 14,006 (1.7) |  | 22,668 (2.7) | 22,668 (2.7) |  | 30,224 (3.5) | 22,998 (2.7) |  |
| 3 | 3,826 (0.5) | 3,826 (0.5) | 0.00 | 9,218 (1.1) | 9,218 (1.1) | 0.00 | 12,835 (1.5) | 9,876 (1.1) | 0.11 |
| 4 | 1,607 (0.2) | 1,607 (0.2) |  | 4,812 (0.6) | 4,812 (0.6) |  | 6,661 (0.8) | 5,427 (0.6) |  |
| 5 | 651 (0.1) | 651 (0.1) |  | 2,649 (0.3) | 2,649 (0.3) |  | 3,599 (0.4) | 3,127 (0.4) |  |
| ≥6 | 240 (0.03) | 240 (0.03) |  | 2,965 (0.4) | 2,965 (0.4) |  | 3,393 (0.4) | 3,402 (0.4) |  |
| Prior infection status<sup>i</sup> |  |  |  |  |  |  |  |  |  |
| No prior infection | 763,818 (94.1) | 763,818 (94.1) |  | 793,066 (93.8) | 793,066 (93.8) |  | 807,899 (93.6) | 807,899 (93.6) |  |
| Prior pre-omicron infection | 46,571 (5.7) | 46,571 (5.7) | 0.00 | 50,741 (6.0) | 50,741 (6.0) | 0.00 | 53,773 (6.2) | 53,773 (6.2) | 0.00 |
| Prior omicron infection | 1,513 (0.2) | 1,513 (0.2) |  | 1,621 (0.2) | 1,621 (0.2) |  | 1,761 (0.2) | 1,761 (0.2) |  |
| Prior pre-omicron & omicron infections | 42 (<0.01) | 42 (<0.01) |  | 41 (<0.01) | 41 (<0.01) |  | 55 (<0.01) | 55 (<0.01) |  |

Abbreviations: IQR, interquartile range; SMD, standardized mean difference.
<sup>a</sup>Cohorts were matched exactly one-to-one by sex, 10-year age group, nationality, and prior infection status. Persons who received their second vaccine dose in a specific calendar week in the two-dose cohort were additionally matched to persons who had a record for a SARS-CoV-2-negative test in that same calendar week in the unvaccinated cohort, to ensure that matched pairs had presence in Qatar over the same time period.
<sup>b</sup>Cohorts were additionally matched by exact types of coexisting conditions.
<sup>c</sup>Cohorts were additionally matched by number of coexisting conditions.
<sup>d</sup>Cohorts were not matched by coexisting conditions.
<sup>e</sup>SMD is the difference in the mean of a covariate between groups divided by the pooled standard deviation. An SMD ≤0.1 indicates adequate matching.
<sup>f</sup>SMD is for the mean difference between groups divided by the pooled standard deviation.
<sup>g</sup>Nationalities were chosen to represent the most populous groups in Qatar.
<sup>b</sup>These comprise up to 156 other nationalities in the different matching analyses.
<sup>i</sup>Ascertained at the start of follow-up.

Estimated effectiveness against infection of the two-dose (primary-series) vaccination relative to no vaccination showed similar values regardless of the matching strategy (Table 2A).

**Table 2.** Hazard ratios for incidence of SARS-CoV-2 infection, severe, critical, or fatal COVID-19, and non-COVID-19 death in the A) two-dose (primary-series) analysis and B) three-dose (booster) analysis using different criteria for matching by coexisting conditions.

|  | Matching by coexisting condition types |  | Matching by number of coexisting conditions |  | No matching by coexisting conditions |  |
| --- | --- | --- | --- | --- | --- | --- |
| A) Two-dose analysis | Two-dose<sup>a,b</sup> | Unvaccinated<sup>a,b</sup> | Two-dose<sup>a,c</sup> | Unvaccinated<sup>a,c</sup> | Two-dose<sup>a,d</sup> | Unvaccinated<sup>a,d</sup> |
| Sample size | 811,944 | 811,944 | 845,469 | 845,469 | 863,488 | 863,488 |
| Number of incident infections | 54,560 | 57,959 | 57,435 | 62,297 | 59,918 | 63,713 |
| Number of severe, critical, or fatal COVID-19 cases | 25 | 619 | 57 | 898 | 72 | 913 |
| Number of non-COVID-19 deaths | 239 | 302 | 285 | 657 | 339 | 715 |
| Total follow-up time (person-weeks) | 47,391,900 | 47,656,092 | 48,496,779 | 48,663,556 | 49,280,631 | 49,499,386 |
| Median follow-up time (IQR; days) | 206 (41-961) | 199 (36-968) | 200 (40-951) | 192 (34-962) | 200 (40-943) | 193 (34-956) |
| **SARS-CoV-2 infection** |  |  |  |  |  |  |
| Incidence rate (10,000 person-weeks; 95% CI) | 11.51 (11.42-11.61) | 12.16 (12.06-12.26) | 11.84 (11.75-11.94) | 12.80 (12.70-12.90) | 12.16 (12.06-12.26) | 12.87 (12.77-12.97) |
| Unadjusted hazard ratio (95% CI) | 0.93 (0.92-0.94) |  | 0.91 (0.90-0.92) |  | 0.93 (0.92-0.94) |  |
| Adjusted hazard ratio (95% CI)<sup>e</sup> | 0.90 (0.89-0.91) |  | 0.88 (0.87-0.89) |  | 0.88 (0.87-0.89) |  |
| Vaccine effectiveness (95% CI) | 10.4 (9.3-11.4) |  | 12.4 (11.4-13.4) |  | 12.0 (11.0-13.0) |  |
| **Severe, critical, or fatal COVID-19** |  |  |  |  |  |  |
| Incidence rate (10,000 person-weeks; 95% CI) | 0.01 (0.004-0.01) | 0.13 (0.12-0.14) | 0.01 (0.01-0.02) | 0.18 (0.17-0.20) | 0.01 (0.01-0.02) | 0.18 (0.17-0.20) |
| Unadjusted hazard ratio (95% CI) | 0.04 (0.03-0.06) |  | 0.06 (0.05-0.08) |  | 0.08 (0.06-0.10) |  |
| Adjusted hazard ratio (95% CI)<sup>e</sup> | 0.04 (0.02-0.05) |  | 0.06 (0.05-0.08) |  | 0.06 (0.05-0.08) |  |
| Vaccine effectiveness (95% CI) | 96.3 (94.5-97.5) |  | 94.2 (92.5-95.6) |  | 94.0 (92.3-95.3) |  |
| **Non-COVID-19 death** |  |  |  |  |  |  |
| Incidence rate (10,000 person-weeks; 95% CI) | 0.05 (0.04-0.06) | 0.06 (0.06-0.07) | 0.06 (0.05-0.07) | 0.14 (0.13-0.15) | 0.07 (0.06-0.08) | 0.14 (0.13-0.16) |
| Unadjusted hazard ratio (95% CI) | 0.79 (0.67-0.93) |  | 0.43 (0.37-0.49) |  | 0.47 (0.41-0.54) |  |
| Adjusted hazard ratio (95% CI)<sup>f</sup> | 0.77 (0.65-0.92) |  | 0.40 (0.35-0.46) |  | 0.38 (0.33-0.43) |  |
| B) Three-dose analysis | Three-dose<sup>g,h</sup> | Two-dose<sup>g,h</sup> | Three-dose<sup>g,c</sup> | Two-dose<sup>g,c</sup> | Three-dose<sup>g,d</sup> | Two-dose<sup>g,d</sup> |
| Sample size | 330,447 | 330,447 | 357,936 | 357,936 | 399,147 | 399,147 |
| Number of incident infections | 27,184 | 35,549 | 31,016 | 41,537 | 35,384 | 47,108 |
| Number of severe, critical, or fatal COVID-19 cases | 6 | 9 | 15 | 43 | 21 | 55 |
| Number of non-COVID-19 deaths | 128 | 145 | 213 | 266 | 313 | 331 |
| Total follow-up time (person-weeks) | 25,023,303 | 24,070,435 | 26,560,057 | 25,333,405 | 29,186,072 | 27,801,284 |
| Median follow-up time (IQR; days) | 730 (65-837) | 720 (49-833) | 724 (59-836) | 707 (44-832) | 716 (56-834) | 698 (43-829) |
| **SARS-CoV-2 infection** |  |  |  |  |  |  |
| Incidence rate (10,000 person-weeks; 95% CI) | 10.86 (10.74-10.99) | 14.77 (14.62-14.92) | 11.68 (11.55-11.81) | 16.40 (16.24-16.55) | 12.12 (12.00-12.25) | 16.94 (16.79-17.10) |
| Unadjusted hazard ratio (95% CI) | 0.74 (0.73-0.75) |  | 0.72 (0.71-0.73) |  | 0.72 (0.71-0.73) |  |
| Adjusted hazard ratio (95% CI)<sup>h</sup> | 0.74 (0.72-0.75) |  | 0.71 (0.70-0.72) |  | 0.71 (0.70-0.72) |  |
| Vaccine effectiveness (95% CI) | 26.4 (25.2-27.6) |  | 28.8 (27.7-29.8) |  | 29.5 (28.5-30.5) |  |
| **Severe, critical, or fatal COVID-19** |  |  |  |  |  |  |
| Incidence rate (10,000 person-weeks; 95% CI) | 0.002 (0.001-0.01) | 0.004 (0.002-0.01) | 0.01 (0.003-0.01) | 0.02 (0.01-0.02) | 0.01 (0.005-0.01) | 0.02 (0.02-0.03) |
| Unadjusted hazard ratio (95% CI) | 0.64 (0.23-1.81) |  | 0.34 (0.19-0.61) |  | 0.37 (0.22-0.61) |  |
| Adjusted hazard ratio (95% CI)<sup>h</sup> | 0.66 (0.23-1.86) |  | 0.36 (0.20-0.66) |  | 0.38 (0.23-0.63) |  |
| Vaccine effectiveness (95% CI) | 34.4 (-46.1-76.8) |  | 63.5 (33.9-79.9) |  | 62.2 (37.0-77.3) |  |
| **Non-COVID-19 death** |  |  |  |  |  |  |
| Incidence rate (10,000 person-weeks; 95% CI) | 0.05 (0.04-0.06) | 0.06 (0.05-0.07) | 0.08 (0.07-0.09) | 0.10 (0.09-0.12) | 0.11 (0.10-0.12) | 0.12 (0.11-0.13) |
| Unadjusted hazard ratio (95% CI) | 0.85 (0.67-1.08) |  | 0.77 (0.64-0.92) |  | 0.90 (0.78-1.06) |  |
| Adjusted hazard ratio (95% CI)<sup>†</sup> | 0.84 (0.66-1.06) |  | 0.69 (0.58-0.83) |  | 0.76 (0.65-0.89) |  |

Abbreviations: CI, confidence interval; COVID-19, coronavirus disease 2019; IQR, interquartile range, SARS-CoV-2, severe acute respiratory syndrome coronavirus 2.
<sup>a</sup>Cohorts were matched exactly one-to-one by sex, 10-year age group, nationality, and prior infection status. Persons who received their second vaccine dose in a specific calendar week in the two-dose cohort were additionally matched to persons who had a record for a SARS-CoV-2-negative test in that same calendar week in the unvaccinated cohort, to ensure that matched pairs had presence in Qatar over the same time period. Persons in the unvaccinated cohort had to be alive, with no documented infection in the 90 days preceding the start of the follow-up, and with no vaccination record at the start of the follow-up.
<sup>b</sup>Cohorts were additionally matched by exact types of coexisting conditions.
<sup>c</sup>Cohorts were additionally matched by number of coexisting conditions.
<sup>d</sup>Cohorts were not matched by coexisting conditions.
<sup>e</sup>Adjusted for sex, 10-year age group, nationality, number of coexisting conditions, prior infection status, calendar week of the second vaccine dose for the two-dose cohort or SARS-CoV-2-negative test for the unvaccinated cohort, and testing rate.
<sup>f</sup>Adjusted for sex, 10-year age group, nationality, number of coexisting conditions, prior infection status, and calendar week of the second vaccine dose for the two-dose cohort or SARS-CoV-2-negative test for the unvaccinated cohort.
<sup>g</sup>Cohorts were matched exactly one-to-one by sex, 10-year age group, nationality, prior infection status, and calendar week of the second vaccine dose. Persons who received their third vaccine dose in a specific calendar week in the three-dose cohort were additionally matched to persons who had a record for a SARS-CoV-2-negative test in that same calendar week in the two-dose cohort, to ensure that matched pairs had presence in Qatar over the same time period. Persons in the two-dose cohort had to be alive, with no documented infection in the 90 days preceding the start of the follow-up, and with no record for vaccination with Dose 3 before the start of the follow-up.
<sup>h</sup>Adjusted for sex, 10-year age group, nationality, number of coexisting conditions, prior infection status, calendar week of the second vaccine dose, and testing rate.
<sup>i</sup>Adjusted for sex, 10-year age group, nationality, number of coexisting conditions, prior infection status, and calendar week of the second vaccine dose.

Effectiveness was 10.4% (95% CI: 9.3-11.4) when matching by coexisting condition types, 12.4% (95% CI: 11.4-13.4) when matching by number of coexisting conditions, and 12.0% (95% CI: 11.0-13.0) with no matching by coexisting conditions.

Similarly, estimated vaccine effectiveness against severe, critical, or fatal COVID-19 showed consistent values across matching strategies (Table 2A). Effectiveness was 96.3% (95% CI: 94.5-97.5) when matching by coexisting condition types, 94.2% (95% CI: 92.5-95.6) when matching by number of coexisting conditions, and 94.0% (95% CI: 92.3-95.3) with no matching by coexisting conditions. Similar results were obtained for individuals clinically vulnerable to severe COVID-19 (Supplementary Table 2).

The time-stratified analysis estimated vaccine effectiveness against severe, critical, or fatal COVID-19 at 98.0% (95% CI: 96.5-98.8) in the first six months of follow-up and 73.2% (95% CI: 47.3-86.4) thereafter when matching by coexisting condition types (Fig. 1). Similar values were observed when matching by number of coexisting conditions or when no matching by coexisting conditions was performed.

**Fig. 1.**
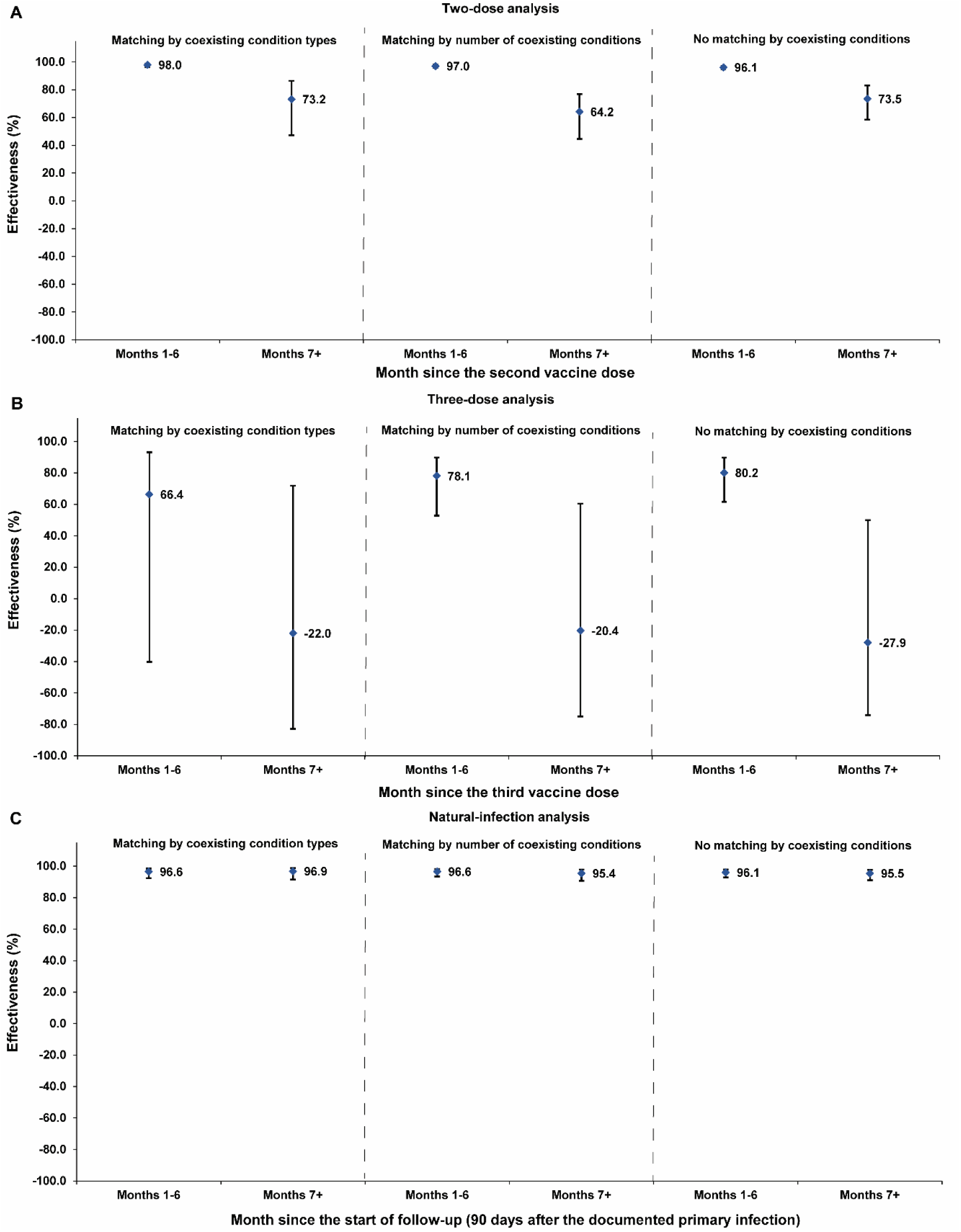
Effectiveness of A) two-dose (primary-series) vaccination, B) three-dose (booster) vaccination, and C) natural infection against severe, critical, or fatal COVID-19 by duration since the start of follow up using different criteria for matching by coexisting conditions.

While vaccine effectiveness estimates were similar regardless of the matching strategy, the aHR for non-COVID-19 death varied, indicating different magnitudes of the healthy vaccinee effect. It was 0.77 (95% CI: 0.65-0.92) when matching by coexisting condition types, 0.40 (95% CI: 0.35-0.46) when matching by number of coexisting conditions, and 0.38 (95% CI: 0.33-0.43) with no matching by coexisting conditions (Table 2A).

### Three-dose analysis

Supplementary Fig. 2 illustrates the matched cohorts’ selection process. Table 3 describes the cohorts’ baseline characteristics. Cohorts comprised 330,447 individuals matched by coexisting condition types, 357,936 matched by number of coexisting conditions, and 399,147 not matched by coexisting conditions.

**Table 3.**
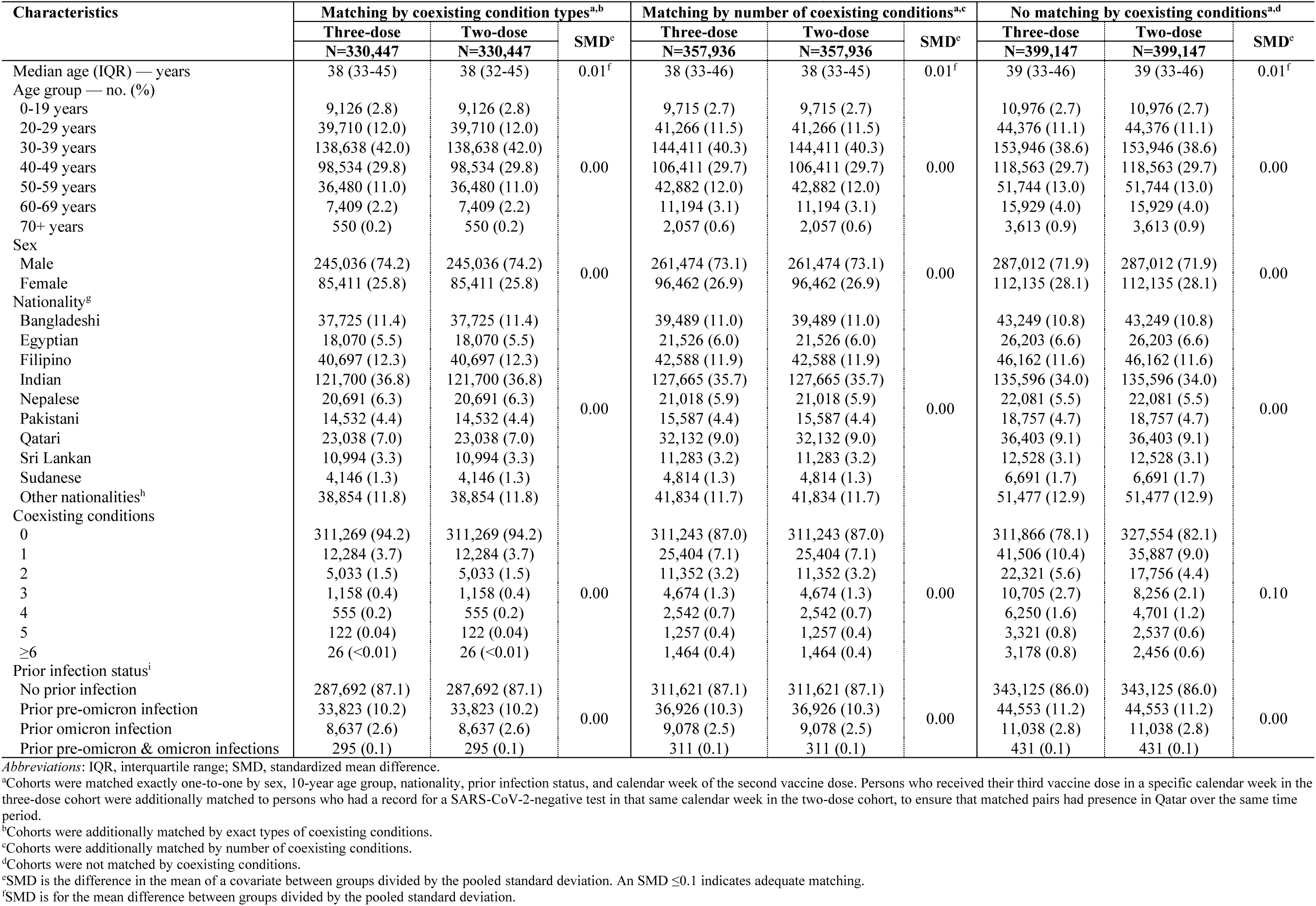
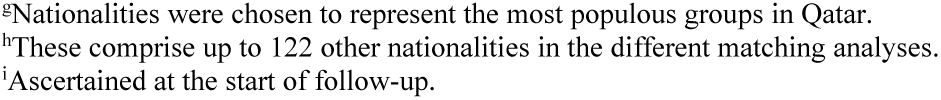
Baseline characteristics of matched cohorts in the three-dose (booster) analysis.

| Characteristics | Matching by coexisting condition types<sup>a,b</sup> |  |  | Matching by number of coexisting conditions<sup>a,c</sup> |  |  | No matching by coexisting conditions<sup>a,d</sup> |  |  |
| --- | --- | --- | --- | --- | --- | --- | --- | --- | --- |
|  | Three-dose | Two-dose | SMD<sup>e</sup> | Three-dose | Two-dose | SMD<sup>e</sup> | Three-dose | Two-dose | SMD<sup>e</sup> |
|  | N=330,447 | N=330,447 |  | N=357,936 | N=357,936 |  | N=399,147 | N=399,147 |  |
| Median age (IQR) — years | 38 (33-45) | 38 (32-45) | 0.01<sup>f</sup> | 38 (33-46) | 38 (33-45) | 0.01<sup>f</sup> | 39 (33-46) | 39 (33-46) | 0.01<sup>f</sup> |
| Age group — no. (%) |  |  |  |  |  |  |  |  |  |
| 0-19 years | 9,126 (2.8) | 9,126 (2.8) |  | 9,715 (2.7) | 9,715 (2.7) |  | 10,976 (2.7) | 10,976 (2.7) |  |
| 20-29 years | 39,710 (12.0) | 39,710 (12.0) |  | 41,266 (11.5) | 41,266 (11.5) |  | 44,376 (11.1) | 44,376 (11.1) |  |
| 30-39 years | 138,638 (42.0) | 138,638 (42.0) |  | 144,411 (40.3) | 144,411 (40.3) |  | 153,946 (38.6) | 153,946 (38.6) |  |
| 40-49 years | 98,534 (29.8) | 98,534 (29.8) | 0.00 | 106,411 (29.7) | 106,411 (29.7) | 0.00 | 118,563 (29.7) | 118,563 (29.7) | 0.00 |
| 50-59 years | 36,480 (11.0) | 36,480 (11.0) |  | 42,882 (12.0) | 42,882 (12.0) |  | 51,744 (13.0) | 51,744 (13.0) |  |
| 60-69 years | 7,409 (2.2) | 7,409 (2.2) |  | 11,194 (3.1) | 11,194 (3.1) |  | 15,929 (4.0) | 15,929 (4.0) |  |
| 70+ years | 550 (0.2) | 550 (0.2) |  | 2,057 (0.6) | 2,057 (0.6) |  | 3,613 (0.9) | 3,613 (0.9) |  |
| Sex |  |  |  |  |  |  |  |  |  |
| Male | 245,036 (74.2) | 245,036 (74.2) | 0.00 | 261,474 (73.1) | 261,474 (73.1) | 0.00 | 287,012 (71.9) | 287,012 (71.9) | 0.00 |
| Female | 85,411 (25.8) | 85,411 (25.8) |  | 96,462 (26.9) | 96,462 (26.9) |  | 112,135 (28.1) | 112,135 (28.1) |  |
| Nationality<sup>g</sup> |  |  |  |  |  |  |  |  |  |
| Bangladeshi | 37,725 (11.4) | 37,725 (11.4) |  | 39,489 (11.0) | 39,489 (11.0) |  | 43,249 (10.8) | 43,249 (10.8) |  |
| Egyptian | 18,070 (5.5) | 18,070 (5.5) |  | 21,526 (6.0) | 21,526 (6.0) |  | 26,203 (6.6) | 26,203 (6.6) |  |
| Filipino | 40,697 (12.3) | 40,697 (12.3) |  | 42,588 (11.9) | 42,588 (11.9) |  | 46,162 (11.6) | 46,162 (11.6) |  |
| Indian | 121,700 (36.8) | 121,700 (36.8) |  | 127,665 (35.7) | 127,665 (35.7) |  | 135,596 (34.0) | 135,596 (34.0) |  |
| Nepalese | 20,691 (6.3) | 20,691 (6.3) | 0.00 | 21,018 (5.9) | 21,018 (5.9) | 0.00 | 22,081 (5.5) | 22,081 (5.5) | 0.00 |
| Pakistani | 14,532 (4.4) | 14,532 (4.4) |  | 15,587 (4.4) | 15,587 (4.4) |  | 18,757 (4.7) | 18,757 (4.7) |  |
| Qatari | 23,038 (7.0) | 23,038 (7.0) |  | 32,132 (9.0) | 32,132 (9.0) |  | 36,403 (9.1) | 36,403 (9.1) |  |
| Sri Lankan | 10,994 (3.3) | 10,994 (3.3) |  | 11,283 (3.2) | 11,283 (3.2) |  | 12,528 (3.1) | 12,528 (3.1) |  |
| Sudanese | 4,146 (1.3) | 4,146 (1.3) |  | 4,814 (1.3) | 4,814 (1.3) |  | 6,691 (1.7) | 6,691 (1.7) |  |
| Other nationalities<sup>h</sup> | 38,854 (11.8) | 38,854 (11.8) |  | 41,834 (11.7) | 41,834 (11.7) |  | 51,477 (12.9) | 51,477 (12.9) |  |
| Coexisting conditions |  |  |  |  |  |  |  |  |  |
| 0 | 311,269 (94.2) | 311,269 (94.2) |  | 311,243 (87.0) | 311,243 (87.0) |  | 311,866 (78.1) | 327,554 (82.1) |  |
| 1 | 12,284 (3.7) | 12,284 (3.7) |  | 25,404 (7.1) | 25,404 (7.1) |  | 41,506 (10.4) | 35,887 (9.0) |  |
| 2 | 5,033 (1.5) | 5,033 (1.5) |  | 11,352 (3.2) | 11,352 (3.2) |  | 22,321 (5.6) | 17,756 (4.4) |  |
| 3 | 1,158 (0.4) | 1,158 (0.4) | 0.00 | 4,674 (1.3) | 4,674 (1.3) | 0.00 | 10,705 (2.7) | 8,256 (2.1) | 0.10 |
| 4 | 555 (0.2) | 555 (0.2) |  | 2,542 (0.7) | 2,542 (0.7) |  | 6,250 (1.6) | 4,701 (1.2) |  |
| 5 | 122 (0.04) | 122 (0.04) |  | 1,257 (0.4) | 1,257 (0.4) |  | 3,321 (0.8) | 2,537 (0.6) |  |
| ≥6 | 26 (<0.01) | 26 (<0.01) |  | 1,464 (0.4) | 1,464 (0.4) |  | 3,178 (0.8) | 2,456 (0.6) |  |
| Prior infection status<sup>i</sup> |  |  |  |  |  |  |  |  |  |
| No prior infection | 287,692 (87.1) | 287,692 (87.1) |  | 311,621 (87.1) | 311,621 (87.1) |  | 343,125 (86.0) | 343,125 (86.0) |  |
| Prior pre-omicron infection | 33,823 (10.2) | 33,823 (10.2) | 0.00 | 36,926 (10.3) | 36,926 (10.3) | 0.00 | 44,553 (11.2) | 44,553 (11.2) | 0.00 |
| Prior omicron infection | 8,637 (2.6) | 8,637 (2.6) |  | 9,078 (2.5) | 9,078 (2.5) |  | 11,038 (2.8) | 11,038 (2.8) |  |
| Prior pre-omicron & omicron infections | 295 (0.1) | 295 (0.1) |  | 311 (0.1) | 311 (0.1) |  | 431 (0.1) | 431 (0.1) |  |

Abbreviations: IQR, interquartile range; SMD, standardized mean difference.
<sup>a</sup>Cohorts were matched exactly one-to-one by sex, 10-year age group, nationality, prior infection status, and calendar week of the second vaccine dose. Persons who received their third vaccine dose in a specific calendar week in the three-dose cohort were additionally matched to persons who had a record for a SARS-CoV-2-negative test in that same calendar week in the two-dose cohort, to ensure that matched pairs had presence in Qatar over the same time period.
<sup>b</sup>Cohorts were additionally matched by exact types of coexisting conditions.
<sup>c</sup>Cohorts were additionally matched by number of coexisting conditions.
<sup>d</sup>Cohorts were not matched by coexisting conditions.
<sup>e</sup>SMD is the difference in the mean of a covariate between groups divided by the pooled standard deviation. An SMD ≤0.1 indicates adequate matching.
<sup>f</sup>SMD is for the mean difference between groups divided by the pooled standard deviation.
<sup>a</sup>Nationalities were chosen to represent the most populous groups in Qatar.
<sup>b</sup>These comprise up to 122 other nationalities in the different matching analyses.
<sup>c</sup>Ascertained at the start of follow-up.

Estimated effectiveness against infection of the three-dose (booster) vaccination relative to the two-dose (primary-series) vaccination showed similar values regardless of the matching strategy (Table 2B). Effectiveness was 26.4% (95% CI: 25.2-27.6) when matching by coexisting condition types, 28.8% (95% CI: 27.7-29.8) when matching by number of coexisting conditions, and 29.5% (95% CI: 28.5-30.5) with no matching by coexisting conditions.

Similarly, estimated vaccine effectiveness against severe, critical, or fatal COVID-19 showed consistent values across matching strategies, but the 95% CIs were wide (Table 2B).

Effectiveness was 34.4% (95% CI: -46.1-76.8) when matching by coexisting condition types, 63.5% (95% CI: 33.9-79.9) when matching by number of coexisting conditions, and 62.2% (95% CI: 37.0-77.3) with no matching by coexisting conditions. Similar results were obtained for individuals clinically vulnerable to severe COVID-19 (Supplementary Table 2).

The time-stratified analysis estimated vaccine effectiveness against severe, critical, or fatal COVID-19 at 66.4% (95% CI: -40.3-93.3) in the first six months of follow-up and 22.0% (95% CI: -82.8-71.8) thereafter when matching by coexisting condition types (Fig. 1). Similar values were observed when matching by number of coexisting conditions or when no matching by coexisting conditions was performed, but the 95% CIs were wide, particularly for effectiveness after the first six months of follow-up.

While vaccine effectiveness estimates were similar regardless of the matching strategy, the aHR for non-COVID-19 death varied, but with somewhat wide 95% CIs. The aHR was 0.84 (95% CI: 0.66-1.06) when matching by coexisting condition types, 0.69 (95% CI: 0.58-0.83) when matching by number of coexisting conditions, and 0.76 (95% CI: 0.65-0.89) with no matching by coexisting conditions (Table 2B).

### Natural-infection analysis

Supplementary Fig. 3 illustrates the matched cohorts’ selection process. Table 4 describes the cohorts’ baseline characteristics. Cohorts comprised 738,564 individuals matched by coexisting condition types, 789,306 matched by number of coexisting conditions, and 816,792 not matched by coexisting conditions.

**Table 4.**
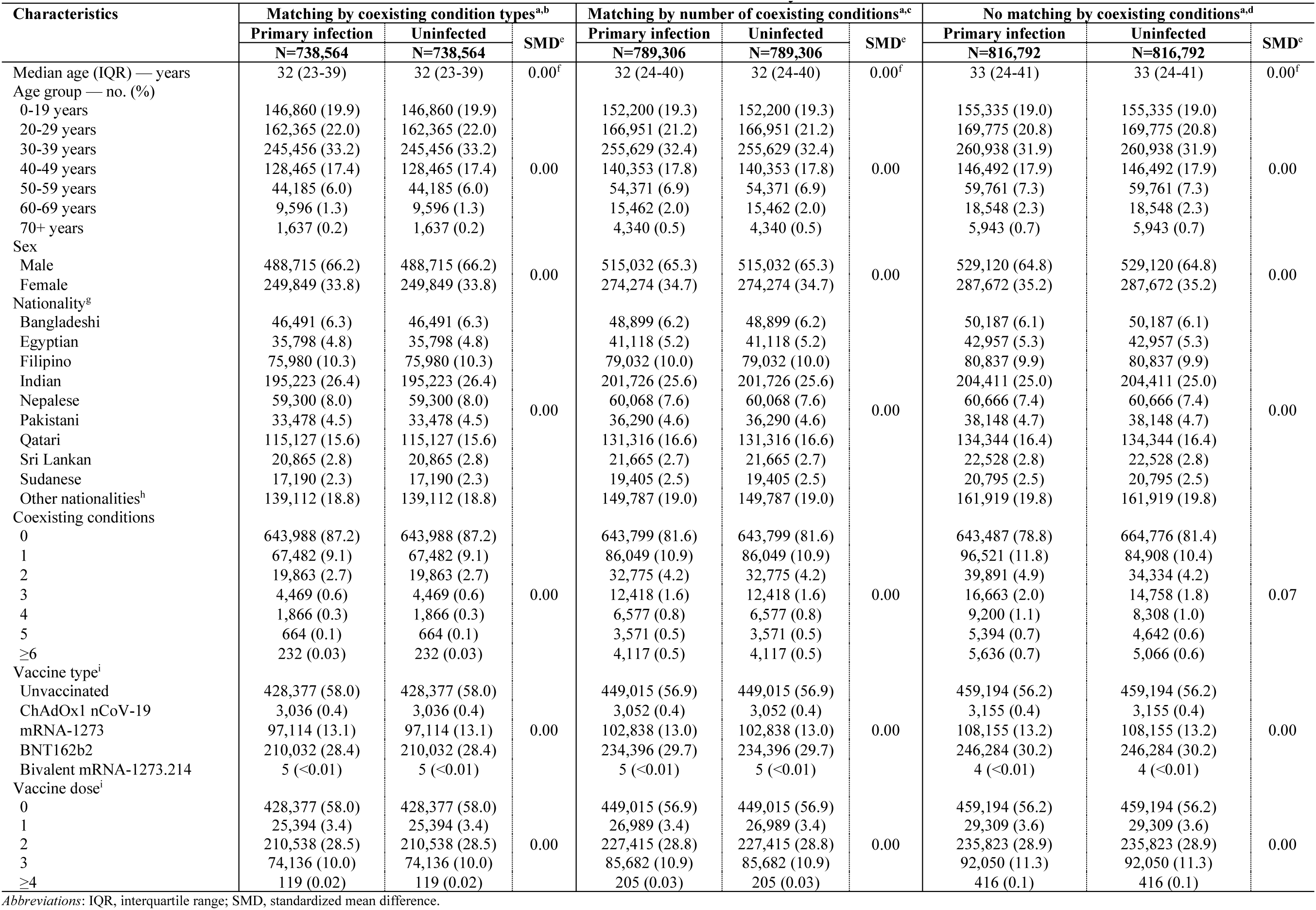
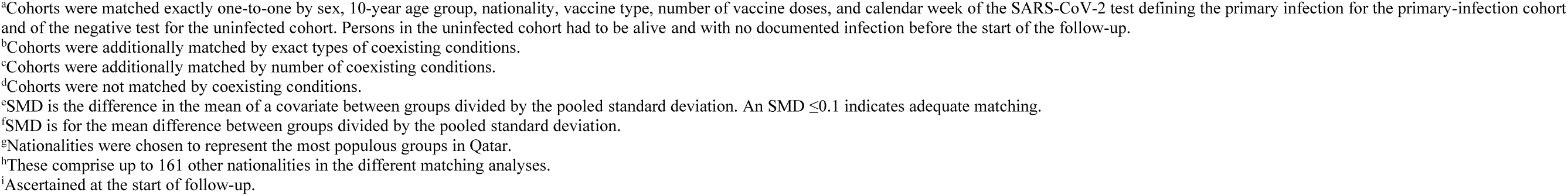
Baseline characteristics of matched cohorts in the natural-infection analysis.

| Characteristics | Matching by coexisting condition types<sup>a,b</sup> |  |  | Matching by number of coexisting conditions<sup>a,c</sup> |  |  | No matching by coexisting conditions<sup>a,d</sup> |  |  |
| --- | --- | --- | --- | --- | --- | --- | --- | --- | --- |
|  | Primary infection | Uninfected | SMD<sup>e</sup> | Primary infection | Uninfected | SMD<sup>e</sup> | Primary infection | Uninfected | SMD<sup>e</sup> |
|  | N=738,564 | N=738,564 |  | N=789,306 | N=789,306 |  | N=816,792 | N=816,792 |  |
| Median age (IQR) — years | 32 (23-39) | 32 (23-39) | 0.00<sup>f</sup> | 32 (24-40) | 32 (24-40) | 0.00<sup>f</sup> | 33 (24-41) | 33 (24-41) | 0.00<sup>f</sup> |
| Age group — no. (%) |  |  |  |  |  |  |  |  |  |
| 0-19 years | 146,860 (19.9) | 146,860 (19.9) |  | 152,200 (19.3) | 152,200 (19.3) |  | 155,335 (19.0) | 155,335 (19.0) |  |
| 20-29 years | 162,365 (22.0) | 162,365 (22.0) |  | 166,951 (21.2) | 166,951 (21.2) |  | 169,775 (20.8) | 169,775 (20.8) |  |
| 30-39 years | 245,456 (33.2) | 245,456 (33.2) |  | 255,629 (32.4) | 255,629 (32.4) |  | 260,938 (31.9) | 260,938 (31.9) |  |
| 40-49 years | 128,465 (17.4) | 128,465 (17.4) | 0.00 | 140,353 (17.8) | 140,353 (17.8) | 0.00 | 146,492 (17.9) | 146,492 (17.9) | 0.00 |
| 50-59 years | 44,185 (6.0) | 44,185 (6.0) |  | 54,371 (6.9) | 54,371 (6.9) |  | 59,761 (7.3) | 59,761 (7.3) |  |
| 60-69 years | 9,596 (1.3) | 9,596 (1.3) |  | 15,462 (2.0) | 15,462 (2.0) |  | 18,548 (2.3) | 18,548 (2.3) |  |
| 70+ years | 1,637 (0.2) | 1,637 (0.2) |  | 4,340 (0.5) | 4,340 (0.5) |  | 5,943 (0.7) | 5,943 (0.7) |  |
| Sex |  |  |  |  |  |  |  |  |  |
| Male | 488,715 (66.2) | 488,715 (66.2) | 0.00 | 515,032 (65.3) | 515,032 (65.3) | 0.00 | 529,120 (64.8) | 529,120 (64.8) | 0.00 |
| Female | 249,849 (33.8) | 249,849 (33.8) |  | 274,274 (34.7) | 274,274 (34.7) |  | 287,672 (35.2) | 287,672 (35.2) |  |
| Nationality<sup>g</sup> |  |  |  |  |  |  |  |  |  |
| Bangladeshi | 46,491 (6.3) | 46,491 (6.3) |  | 48,899 (6.2) | 48,899 (6.2) |  | 50,187 (6.1) | 50,187 (6.1) |  |
| Egyptian | 35,798 (4.8) | 35,798 (4.8) |  | 41,118 (5.2) | 41,118 (5.2) |  | 42,957 (5.3) | 42,957 (5.3) |  |
| Filipino | 75,980 (10.3) | 75,980 (10.3) |  | 79,032 (10.0) | 79,032 (10.0) |  | 80,837 (9.9) | 80,837 (9.9) |  |
| Indian | 195,223 (26.4) | 195,223 (26.4) |  | 201,726 (25.6) | 201,726 (25.6) |  | 204,411 (25.0) | 204,411 (25.0) |  |
| Nepalese | 59,300 (8.0) | 59,300 (8.0) | 0.00 | 60,068 (7.6) | 60,068 (7.6) | 0.00 | 60,666 (7.4) | 60,666 (7.4) | 0.00 |
| Pakistani | 33,478 (4.5) | 33,478 (4.5) |  | 36,290 (4.6) | 36,290 (4.6) |  | 38,148 (4.7) | 38,148 (4.7) |  |
| Qatari | 115,127 (15.6) | 115,127 (15.6) |  | 131,316 (16.6) | 131,316 (16.6) |  | 134,344 (16.4) | 134,344 (16.4) |  |
| Sri Lankan | 20,865 (2.8) | 20,865 (2.8) |  | 21,665 (2.7) | 21,665 (2.7) |  | 22,528 (2.8) | 22,528 (2.8) |  |
| Sudanese | 17,190 (2.3) | 17,190 (2.3) |  | 19,405 (2.5) | 19,405 (2.5) |  | 20,795 (2.5) | 20,795 (2.5) |  |
| Other nationalities<sup>h</sup> | 139,112 (18.8) | 139,112 (18.8) |  | 149,787 (19.0) | 149,787 (19.0) |  | 161,919 (19.8) | 161,919 (19.8) |  |
| Coexisting conditions |  |  |  |  |  |  |  |  |  |
| 0 | 643,988 (87.2) | 643,988 (87.2) |  | 643,799 (81.6) | 643,799 (81.6) |  | 643,487 (78.8) | 664,776 (81.4) |  |
| 1 | 67,482 (9.1) | 67,482 (9.1) |  | 86,049 (10.9) | 86,049 (10.9) |  | 96,521 (11.8) | 84,908 (10.4) |  |
| 2 | 19,863 (2.7) | 19,863 (2.7) | 0.00 | 32,775 (4.2) | 32,775 (4.2) | 0.00 | 39,891 (4.9) | 34,334 (4.2) | 0.07 |
| 3 | 4,469 (0.6) | 4,469 (0.6) |  | 12,418 (1.6) | 12,418 (1.6) |  | 16,663 (2.0) | 14,758 (1.8) |  |
| 4 | 1,866 (0.3) | 1,866 (0.3) |  | 6,577 (0.8) | 6,577 (0.8) |  | 9,200 (1.1) | 8,308 (1.0) |  |
| 5 | 664 (0.1) | 664 (0.1) |  | 3,571 (0.5) | 3,571 (0.5) |  | 5,394 (0.7) | 4,642 (0.6) |  |
| ≥6 | 232 (0.03) | 232 (0.03) |  | 4,117 (0.5) | 4,117 (0.5) |  | 5,636 (0.7) | 5,066 (0.6) |  |
| Vaccine type<sup>i</sup> |  |  |  |  |  |  |  |  |  |
| Unvaccinated | 428,377 (58.0) | 428,377 (58.0) |  | 449,015 (56.9) | 449,015 (56.9) |  | 459,194 (56.2) | 459,194 (56.2) |  |
| ChAdOx1 nCoV-19 | 3,036 (0.4) | 3,036 (0.4) | 0.00 | 3,052 (0.4) | 3,052 (0.4) | 0.00 | 3,155 (0.4) | 3,155 (0.4) | 0.00 |
| mRNA-1273 | 97,114 (13.1) | 97,114 (13.1) |  | 102,838 (13.0) | 102,838 (13.0) |  | 108,155 (13.2) | 108,155 (13.2) |  |
| BNT162b2 | 210,032 (28.4) | 210,032 (28.4) |  | 234,396 (29.7) | 234,396 (29.7) |  | 246,284 (30.2) | 246,284 (30.2) |  |
| Bivalent mRNA-1273.214 | 5 (<0.01) | 5 (<0.01) |  | 5 (<0.01) | 5 (<0.01) |  | 4 (<0.01) | 4 (<0.01) |  |
| Vaccine dose<sup>i</sup> |  |  |  |  |  |  |  |  |  |
| 0 | 428,377 (58.0) | 428,377 (58.0) |  | 449,015 (56.9) | 449,015 (56.9) |  | 459,194 (56.2) | 459,194 (56.2) |  |
| 1 | 25,394 (3.4) | 25,394 (3.4) | 0.00 | 26,989 (3.4) | 26,989 (3.4) | 0.00 | 29,309 (3.6) | 29,309 (3.6) | 0.00 |
| 2 | 210,538 (28.5) | 210,538 (28.5) |  | 227,415 (28.8) | 227,415 (28.8) |  | 235,823 (28.9) | 235,823 (28.9) |  |
| 3 | 74,136 (10.0) | 74,136 (10.0) |  | 85,682 (10.9) | 85,682 (10.9) |  | 92,050 (11.3) | 92,050 (11.3) |  |
| ≥4 | 119 (0.02) | 119 (0.02) |  | 205 (0.03) | 205 (0.03) |  | 416 (0.1) | 416 (0.1) |  |

Abbreviations: IQR, interquartile range; SMD, standardized mean difference.
<sup>a</sup>Cohorts were matched exactly one-to-one by sex, 10-year age group, nationality, vaccine type, number of vaccine doses, and calendar week of the SARS-CoV-2 test defining the primary infection for the primary-infection cohort and of the negative test for the uninfected cohort. Persons in the uninfected cohort had to be alive and with no documented infection before the start of the follow-up.
<sup>b</sup>Cohorts were additionally matched by exact types of coexisting conditions.
<sup>c</sup>Cohorts were additionally matched by number of coexisting conditions.
<sup>d</sup>Cohorts were not matched by coexisting conditions.
<sup>e</sup>SMD is the difference in the mean of a covariate between groups divided by the pooled standard deviation. An $SMD \leq 0.1$ indicates adequate matching.
<sup>f</sup>SMD is for the mean difference between groups divided by the pooled standard deviation.
<sup>g</sup>Nationalities were chosen to represent the most populous groups in Qatar.
<sup>h</sup>These comprise up to 161 other nationalities in the different matching analyses.
<sup>i</sup>Ascertained at the start of follow-up.

Estimated effectiveness of natural infection against reinfection showed similar values regardless of the matching strategy (Table 5). Effectiveness was 50.4% (95% CI: 49.7-51.1) when matching by coexisting condition types, 50.5% (95% CI: 49.8-51.1) when matching by number of coexisting conditions, and 50.2% (95% CI: 49.6-50.8) with no matching by coexisting conditions.

**Table 5.** Hazard ratios for incidence of SARS-CoV-2 infection, severe, critical, or fatal COVID-19, and non-COVID-19 death in the natural-infection analysis using different criteria for matching by coexisting conditions.

| Natural-infection analysis | Matching by coexisting condition types |  | Matching by number of coexisting conditions |  | No matching by coexisting conditions |  |
| --- | --- | --- | --- | --- | --- | --- |
|  | Primary infection<sup>a,b</sup> | Uninfected<sup>a,b</sup> | Primary infection<sup>a,c</sup> | Uninfected<sup>a,c</sup> | Primary infection<sup>a,d</sup> | Uninfected<sup>a,d</sup> |
| Sample size | 738,564 | 738,564 | 789,306 | 789,306 | 816,792 | 816,792 |
| Number of incident infections | 32,451 | 60,672 | 36,061 | 67,106 | 37,793 | 69,118 |
| Number of severe, critical, or fatal COVID-19 cases | 10 | 299 | 18 | 437 | 21 | 464 |
| Number of non-COVID-19 deaths | 282 | 312 | 571 | 695 | 729 | 785 |
| Total follow-up time (person-weeks) | 50,244,439 | 48,056,032 | 53,154,467 | 50,741,522 | 54,644,276 | 52,193,379 |
| Median follow-up time (IQR; days) | 495 (159-767) | 430 (140-765) | 485 (155-766) | 402 (136-765) | 477 (153-766) | 388 (133-764) |
| **SARS-CoV-2 infection** |  |  |  |  |  |  |
| Incidence rate (10,000 person-weeks; 95% CI) | 6.46 (6.39-6.53) | 12.63 (12.53-12.73) | 6.78 (6.71-6.85) | 13.23 (13.13-13.33) | 6.92 (6.85-6.99) | 13.24 (13.14-13.34) |
| Unadjusted hazard ratio (95% CI) | 0.51 (0.51-0.52) |  | 0.52 (0.51-0.52) |  | 0.53 (0.52-0.53) |  |
| Adjusted hazard ratio (95% CI)<sup>e</sup> | 0.50 (0.49-0.50) |  | 0.50 (0.49-0.50) |  | 0.50 (0.49-0.50) |  |
| Natural-infection effectiveness (95% CI) | 50.4 (49.7-51.1) |  | 50.5 (49.8-51.1) |  | 50.2 (49.6-50.8) |  |
| **Severe, critical, or fatal COVID-19** |  |  |  |  |  |  |
| Incidence rate (10,000 person-weeks; 95% CI) | 0.002 (0.001-0.004) | 0.06 (0.06-0.07) | 0.003 (0.002-0.01) | 0.09 (0.08-0.09) | 0.004 (0.003-0.01) | 0.09 (0.08-0.10) |
| Unadjusted hazard ratio (95% CI) | 0.03 (0.02-0.06) |  | 0.04 (0.02-0.06) |  | 0.04 (0.03-0.07) |  |
| Adjusted hazard ratio (95% CI)<sup>e</sup> | 0.03 (0.02-0.06) |  | 0.04 (0.02-0.06) |  | 0.04 (0.03-0.06) |  |
| Natural-infection effectiveness (95% CI) | 96.7 (93.8-98.2) |  | 96.2 (93.8-97.6) |  | 95.9 (93.5-97.4) |  |
| **Non-COVID-19 death** |  |  |  |  |  |  |
| Incidence rate (10,000 person-weeks; 95% CI) | 0.06 (0.05-0.06) | 0.06 (0.06-0.07) | 0.11 (0.10-0.12) | 0.14 (0.13-0.15) | 0.13 (0.12-0.14) | 0.15 (0.14-0.16) |
| Unadjusted hazard ratio (95% CI) | 0.87 (0.74-1.02) |  | 0.79 (0.71-0.88) |  | 0.89 (0.81-0.99) |  |
| Adjusted hazard ratio (95% CI)<sup>f</sup> | 0.86 (0.73-1.01) |  | 0.75 (0.67-0.84) |  | 0.81 (0.73-0.90) |  |

Abbreviations: CI, confidence interval; COVID-19, coronavirus disease 2019; SARS-CoV-2, severe acute respiratory syndrome coronavirus 2.
<sup>a</sup>Cohorts were matched exactly one-to-one by sex, 10-year age group, nationality, prior vaccine type, number of vaccine doses, and calendar week of the SARS-CoV-2 test defining the primary infection for the primary-infection cohort and of the negative test for the uninfected cohort. Persons in the uninfected cohort had to be alive and with no documented infection before the start of the follow-up
<sup>b</sup>Cohorts were additionally matched by exact types of coexisting conditions.
<sup>c</sup>Cohorts were additionally matched by number of coexisting conditions.
<sup>d</sup>Cohorts were not matched by coexisting conditions.
<sup>e</sup>Adjusted for sex, 10-year age group, nationality, number of coexisting conditions, prior vaccine type, number of vaccine doses, calendar week of the SARS-CoV-2 test defining the primary infection for the primary-infection cohort and of the negative test for the uninfected cohort, and testing rate.
<sup>f</sup>Adjusted for sex, 10-year age group, nationality, number of coexisting conditions, prior vaccine type, number of vaccine doses, and calendar week of the SARS-CoV-2 test defining the primary infection for the primary-infection cohort and of the negative test for the uninfected cohort.

Similarly, estimated effectiveness of natural infection against severe, critical, or fatal COVID-19 upon reinfection showed consistent values across matching strategies (Table 5). Effectiveness was 96.7% (95% CI: 93.8-98.2) when matching by coexisting condition types, 96.2% (95% CI: 93.8-97.6) when matching by number of coexisting conditions, and 95.9% (95% CI: 93.5-97.4) with no matching by coexisting conditions. The time-stratified analysis estimated this effectiveness to remain above 95% throughout the follow-up period, regardless of the matching criteria used for coexisting conditions (Fig. 1). Similar results were obtained for individuals clinically vulnerable to severe COVID-19 (Supplementary Table 2).

Unlike in the vaccine two-dose analysis, the aHR for non-COVID-19 death did not appreciably vary by matching strategy. The aHR was 0.86 (95% CI: 0.73-1.01) when matching by coexisting condition types, 0.75 (95% CI: 0.67-0.84) when matching by number of coexisting conditions, and 0.81 (95% CI: 0.73-0.90) with no matching by coexisting conditions (Table 5).

## Discussion

The results demonstrated the presence of the healthy vaccinee effect, with vaccinated individuals exhibiting better health in terms of a lower risk of non-COVID-19 death compared to unvaccinated individuals. This effect, consistent with our earlier observations[9], persisted across matching strategies but was minimized when matching by exact types of coexisting conditions.

This outcome is expected, as this matching reduces the imbalance in health status between vaccinated and unvaccinated individuals. The healthy vaccinee effect was more pronounced in the two-dose analysis compared to the three-dose analysis. This is anticipated because the two-dose analysis compared vaccinated versus unvaccinated individuals, whereas the three-dose analysis compared two vaccinated groups, differing only in their willingness to receive a third dose.

Strikingly, the imbalance in health status between vaccinated and unvaccinated persons did not introduce a healthy vaccinee bias in vaccine effectiveness estimates, whether against infection or, more importantly, against severe, critical, or fatal COVID-19. Effectiveness estimates remained consistently similar across the different matching strategies for coexisting conditions, even though the healthy vaccinee effect varied in magnitude across these strategies. This consistency held true shortly after vaccination, in the long term, and among individuals clinically vulnerable to severe COVID-19, despite the healthy vaccinee effect being more pronounced in the first six months after vaccination and among those clinically vulnerable to severe COVID-19 [1, 5, 9].

These findings underscore the critical distinction between the healthy vaccinee effect and the healthy vaccinee bias. While the healthy vaccinee effect reflects inherent differences in health status between vaccinated and unvaccinated individuals in real-world conditions, the healthy vaccinee bias occurs only when this disparity distorts vaccine effectiveness estimates. In this study, such bias was not observed. Despite varying magnitudes of the healthy vaccinee effect across the different matching strategies, vaccine effectiveness estimates remained consistently similar.

The absence of a healthy vaccinee bias may stem from the rigorous assessment of outcomes, employing laboratory-confirmed infections and stringent WHO criteria for defining COVID-19 severity [27], criticality [27], and fatality [26]. This emphasizes the importance of selecting specific and well-defined outcomes in vaccine effectiveness studies, rather than relying on nonspecific or proxy measures such as all-cause mortality or general respiratory hospitalizations [1, 3, 5, 39]. The observed consistency in vaccine effectiveness across the different matching strategies for coexisting conditions may also suggest that COVID-19 vaccines provide largely comparable protection regardless of individual health conditions, thereby mitigating the impact of the healthy vaccinee effect.

While the healthy vaccinee effect was evident in the vaccination analyses, the corresponding effect in the natural-infection analysis was minimal. Non-COVID-19 death rates were largely comparable between the primary-infection and uninfected cohorts, and matching by coexisting condition types balanced health status. This contrasts with the vaccination analyses, where an imbalance in health status persisted even after matching by coexisting condition types. This finding may be attributed to the voluntary nature of vaccination, which can lead to selection bias in health status, whereas infection is generally a random event with less influence from individual choices.

The study presents findings from this stage of the pandemic that align with earlier observations. Vaccine effectiveness against infection from the primary-series or booster vaccination was modest over long follow-up durations, since this type of protection diminishes rapidly within a few months [10, 40–42]. However, vaccine effectiveness against severe, critical, or fatal COVID-19 remained high, although it waned over time at a slower rate. In contrast, natural infection provided about 50% protection against reinfection throughout the follow-up period, and showed no evidence of waning against severe, critical, or fatal COVID-19 at reinfection. These findings confirm earlier results, including those observed in this specific population [10–12, 17, 19, 22, 23, 30, 40–48], and suggest that the protection conferred by natural infection is both stronger and more durable than that provided by vaccination, perhaps due to the distinct roles of mucosal immunity in these two immune response mechanisms [23, 49, 50].

This study has limitations. The relatively small number of severe, critical, or fatal COVID-19 cases among vaccinated individuals resulted in wide CIs for some estimates in the three-dose analysis. Coexisting conditions were identified using electronic health records, but documented cases may not capture all instances of coexisting conditions. Similarly, documented SARS-CoV-2 infections do not account for all infections that have occurred in the population. However, since under-ascertainment of these outcomes may have affected the study cohorts similarly, their impact on the study estimates—based on relative comparisons between the cohorts—may be mitigated.

Documented COVID-19 deaths may not capture all fatalities attributed to the virus [51, 52], potentially leading to misclassification bias. However, the relatively very low COVID-19 mortality rate in Qatar’s young and working-age population [16, 18, 20, 53], along with previous findings indicating minimal undocumented COVID-19 deaths [18, 20], suggests that this bias likely had a limited impact on the study’s findings.

Considering Qatar’s relatively young population, the study findings may not be generalizable to countries with a larger elderly or comorbid population [16]. However, the results remained consistent in the subgroup analysis that included only individuals aged over 50 years or those with two or more coexisting conditions. As an observational study, the cohorts were neither blinded nor randomized, leaving open the possibility of unmeasured or uncontrolled confounders. Although rigorous matching was performed for factors affecting the risks of infection and severe forms of COVID-19 [16, 54–57], matching was not possible for certain factors, such as geography or occupation, due to data unavailability. Nonetheless, Qatar’s status as a city-state with a broadly distributed infection incidence across neighborhoods, combined with evidence that nationality, age, and sex serve as effective proxies for socioeconomic status and occupation in this country [16, 54–57], suggests that these limitations may not have impacted the study results. The matching procedure used in this study has also been evaluated in previous studies with different epidemiologic designs and using control groups to test for null effects [10, 14, 15, 41, 58]. These prior studies demonstrated that this procedure effectively balances differences to estimate vaccine effectiveness [10, 14, 15, 41, 58].

The study has strengths. It included Qatar’s entire population, featuring sizable cohorts representing diverse national backgrounds. The availability of an integrated digital health information platform provided access to extensive and validated databases, allowing for rigorous matching on various confounding factors such as specific coexisting conditions and previous vaccination and infection statuses. Ascertainment of severe, critical, and fatal COVID-19 cases was meticulously conducted following stringent WHO criteria [26, 27]. The study also examined effects in two independent vaccination analyses: one for the primary series and one for the third booster dose.

In conclusion, while the study found evidence of the healthy vaccinee effect—where vaccinated individuals had better overall health status than unvaccinated individuals—this imbalance did not bias the estimated vaccine effectiveness against infection or severe, critical, and fatal COVID-19, whether shortly after vaccination, in the long term, or among individuals clinically vulnerable to severe COVID-19. The absence of bias in vaccine effectiveness estimates may be attributed to the rigorous and specific assessment of outcomes, with severe COVID-19 cases adhering to WHO criteria. These results highlight the importance of using specific, well-defined outcomes in vaccine effectiveness studies.

## Author Contributions

H.C. co-designed the study, performed the statistical analyses, and co-wrote the first draft of the article. L.J.A. conceived and co-designed the study, led the statistical analyses, and co-wrote the first draft of the article. H.C. and L.J.A. accessed and verified all the data. P.V.C. designed mass PCR testing to allow routine capture of variants and conducted viral genome sequencing. P.T. and M.R.H. designed and conducted multiplex, RT-qPCR variant screening and viral genome sequencing. H.M.Y. and A.A.AT. conducted viral genome sequencing. All authors contributed to data collection and acquisition, database development, discussion and interpretation of the results, and to the writing of the article. All authors have read and approved the final manuscript.

## Acknowledgments

The authors acknowledge the many dedicated individuals at Hamad Medical Corporation, the Ministry of Public Health, the Primary Health Care Corporation, Qatar Biobank, Sidra Medicine, and Weill Cornell Medicine-Qatar for their diligent efforts and contributions to make this study possible. The authors are also grateful for institutional salary support from the Biomedical Research Program and the Biostatistics, Epidemiology, and Biomathematics Research Core, both at Weill Cornell Medicine-Qatar, as well as for institutional salary support provided by the Ministry of Public Health, Hamad Medical Corporation, and Sidra Medicine. HC gratefully acknowledges salary support from the Junior Faculty Transition to Independence Program at Weill Cornell Medicine–Qatar and L’Oréal-UNESCO For Women In Science Middle East Regional Young Talents Program. The authors are also grateful for the Qatar Genome Programme and Qatar University Biomedical Research Center for institutional support for the reagents needed for the viral genome sequencing. The funders of the study had no role in study design, data collection, data analysis, data interpretation, or writing of the article. Statements made herein are solely the responsibility of the authors.

## Declarations Ethics approval

The institutional review boards at Hamad Medical Corporation and Weill Cornell Medicine– Qatar approved this retrospective study with a waiver of informed consent.

## Data availability

The dataset of this study is a property of the Qatar Ministry of Public Health that was provided to the researchers through a restricted-access agreement that prevents sharing the dataset with a third party or publicly. The data are available under restricted access for preservation of confidentiality of patient data. Access can be obtained through a direct application for data access to His Excellency the Minister of Public Health (https://emsfsa.moph.gov.qa/en/Pages/eservices.aspx). The raw data are protected and are not available due to data privacy laws. Requests for access are assessed by the Ministry of Public

Health in Qatar, and approval is granted at its discretion. In compliance with data privacy laws and the data-sharing agreement with the Ministry of Public Health in Qatar, no datasets, whether raw or de-identified, can be publicly released by the researchers. Aggregate data are available within the paper and its supplementary information.

## Use of Artificial Intelligence

ChatGPT was used for grammar verification and phrasing refinement. The authors thoroughly reviewed and edited the text, taking full responsibility for its accuracy and quality.

## Supplementary Section 1. Study population and data sources

Qatar’s national and universal public healthcare system uses the Cerner-system advanced digital health platform to track all electronic health record encounters of each individual in the country, including all citizens and residents registered in the national and universal public healthcare system. Registration in the public healthcare system is mandatory for citizens and residents.

The databases analyzed in this study are data-extract downloads from the Cerner-system that have been implemented on a regular weekly schedule since the onset of pandemic by the Business Intelligence Unit at Hamad Medical Corporation (HMC). HMC is the national public healthcare provider in Qatar. At every download all severe acute respiratory syndrome coronavirus 2 (SARS-CoV-2) tests, coronavirus disease 2019 (COVID-19) vaccinations, hospitalizations related to COVID-19, and all death records regardless of cause are provided to the authors through .csv files. These databases have been analyzed throughout the pandemic not only for study-related purposes, but also to provide policymakers with summary data and analytics to inform the national response.

Every health encounter in the Cerner-system is linked to an individual through the HMC Number, which serves as a unique identifier that links all records for this individual at the national level. Databases were merged and analyzed using the HMC Number to link all records pertaining to testing, vaccinations, hospitalizations, and deaths. All deaths in Qatar are recorded by the public healthcare system. COVID-19-related healthcare was provided exclusively in the public healthcare system. COVID-19 vaccination was also provided only through the public healthcare system. These health records were tracked throughout the COVID-19 pandemic using the Cerner system. This system has been implemented in 2013, before the onset of the pandemic. This pre-established system ensured that we had access to comprehensive health records related to this study for both citizens and residents throughout the entire pandemic, allowing us to follow each person over time.

Demographic details for every HMC Number (individual) such as sex, age, and nationality are collected upon issuing of the universal health card, based on the Qatar Identity Card, which is a mandatory requirement by the Ministry of Interior to every citizen and resident in the country. Data extraction from the Qatar Identity Card to the digital health platform is performed electronically through scanning techniques.

All SARS-CoV-2 testing in any facility in Qatar is tracked nationally in one database, the national testing database. This database covers all testing throughout the country, whether in public or private facilities. Every polymerase chain reaction (PCR) test and a proportion of the facility-based rapid antigen tests conducted in Qatar, regardless of location or setting, are classified on the basis of symptoms and the reason for testing, such as the presence of clinical symptoms, contact tracing, participation in surveys or random testing campaigns, individual requests for testing, routine healthcare testing, pre-travel requirements, at the point of entry into the country, or any other relevant reasons for testing.

Before November 1, 2022, SARS-CoV-2 testing in Qatar was performed extensively with about 5% of the population tested every week [1]. Based on the distribution of the reason for testing up to November 1, 2022, most of the tests in Qatar were conducted for routine reasons, such as travel-related purposes, and about 75% of infections were diagnosed not because of presence of symptoms [1, 2]. Starting from November 1, 2022, testing for SARS-CoV-2 was substantially reduced with <1% of the population tested every week [2]. This study factored all SARS-CoV-2- related testing included in the national testing database over the duration of follow-up.

December 19, 2021 marked the onset of the omicron wave in Qatar [1]. The first omicron wave that reached its peak in January of 2022 was massive and strained the testing capacity in the country [1, 3–5]. To alleviate the burden on PCR testing, rapid antigen testing was rapidly introduced. The swift change in testing policy precluded incorporating reason for testing for a number of rapid antigen tests. While the reason for testing is documented for all PCR tests, it is not uniformly available for all rapid antigen tests. However, all medically supervised rapid antigen tests were captured by the national integrated digital health platform since January 5, 2022.

Rapid antigen test kits are accessible for purchase at pharmacies in Qatar, but results of home-based testing are neither reported nor documented in the national databases. Since SARS-CoV-2- test outcomes were linked to specific public health measures, restrictions, and privileges, testing policy and guidelines stress facility-based testing as the core testing mechanism in the population. While facility-based testing is provided free of charge or at low subsidized costs, depending on the reason for testing, home-based rapid antigen testing is de-emphasized and not supported as part of national policy.

Qatar launched its COVID-19 vaccination program in December 2020, employing mRNA vaccines and prioritizing individuals based on coexisting conditions and age criteria [2, 6]. COVID-19 vaccination was provided free of charge, regardless of citizenship or residency status, and was nationally tracked [2, 6].

Qatar has unusually young, diverse demographics, in that only 9% of its residents are ≥50 years of age, and 89% are expatriates from over 150 countries [2, 7]. Further descriptions of the study population and these national databases were reported previously [1, 2, 5, 8–11].

## Supplementary Section 2. Laboratory methods and variant ascertainment Real-time reverse-transcription polymerase chain reaction testing

Nasopharyngeal and/or oropharyngeal swabs were collected for PCR testing and placed in Universal Transport Medium (UTM). Aliquots of UTM were: 1) extracted on KingFisher Flex (Thermo Fisher Scientific, USA), MGISP-960 (MGI, China), or ExiPrep 96 Lite (Bioneer, South Korea) followed by testing with real-time reverse-transcription PCR (RT-qPCR) using TaqPath COVID-19 Combo Kits (Thermo Fisher Scientific, USA) on an ABI 7500 FAST (Thermo Fisher Scientific, USA); 2) tested directly on the Cepheid GeneXpert system using the Xpert Xpress SARS-CoV-2 (Cepheid, USA); or 3) loaded directly into a Roche cobas 6800 system and assayed with the cobas SARS-CoV-2 Test (Roche, Switzerland). The first assay targets the viral S, N, and ORF1ab gene regions. The second targets the viral N and E-gene regions, and the third targets the ORF1ab and E-gene regions.

All PCR testing was conducted at the Hamad Medical Corporation Central Laboratory or Sidra Medicine Laboratory, following standardized protocols.

## Rapid antigen testing

SARS-CoV-2 antigen tests were performed on nasopharyngeal swabs using one of the following lateral flow antigen tests: Panbio COVID-19 Ag Rapid Test Device (Abbott, USA); SARS-CoV- 2 Rapid Antigen Test (Roche, Switzerland); Standard Q COVID-19 Antigen Test (SD Biosensor, Korea); or CareStart COVID-19 Antigen Test (Access Bio, USA). All antigen tests were performed at point-of-care according to each manufacturer’s instructions, at public or private hospitals and clinics throughout Qatar, with prior authorization and training by the Ministry of Public Health (MOPH). Antigen test results were electronically reported to the MOPH in real time using the Antigen Test Management System which is integrated with the national COVID- 19 database.

## Classification of infections by variant type

Surveillance for SARS-CoV-2 variants in Qatar is based on viral genome sequencing and multiplex reverse transcription quantitative PCR (RT-qPCR) variant screening [12] of weekly collected random positive clinical samples [2, 13–17], complemented by deep sequencing of wastewater samples [15, 18, 19]. Further details on the viral genome sequencing and multiplex RT-qPCR variant screening throughout the SARS-CoV-2 waves in Qatar can be found in previous publications [1, 2, 4, 9, 13–17, 20–25].

## Supplementary Section 3. Classification of coexisting conditions

Coexisting conditions were ascertained and classified based on the ICD-10 codes for the conditions, as recorded in the electronic health record encounters of each individual in the Cerner-system national database that includes all citizens and residents registered in the national and universal public healthcare system. The public healthcare system provides healthcare to the entire resident population of Qatar free of charge or at heavily subsidized costs, including prescription drugs. With the mass expansion of this sector in recent years, facilities have been built to cater to specific needs of subpopulations. For example, tens of facilities have been built, including clinics and hospitals, in localities with high density of craft and manual workers[26].

All encounters for each individual were analyzed to determine the coexisting-condition classification for that individual. The Cerner-system national database includes encounters starting from 2013, when this system was launched in Qatar. Any individual who had at least one encounter with a specific coexisting-condition diagnosis since 2013 was classified as having that coexisting condition. Individuals who do not have records of coexisting-condition encounters in the public healthcare system were classified as having no coexisting conditions.

The classification of coexisting conditions spanned the following conditions: 1) Behchet’s disease, 2) cancer, 3) cardiovascular diseases, 4) infectious and parasitic diseases, 5) Chron’s disease, 6) chronic kidney disease (CKD), 7) chronic liver disease (CLD), 8) chronic lung disease, 9) congenital malformations, deformations and chromosomal abnormalities, 10) diseases of the blood and blood-forming organs and certain disorders involving the immune mechanism, 11) diseases of the ear and mastoid process, 12) deep vein thrombosis (DVT), 13) dermatitis, 14) diabetes mellitus, 15) diseases of the circulatory system, 16) diseases of the digestive system, 17) diseases of the eye and adnex, 18) diseases of the genitourinary system, 19) diseases of the musculoskeletal system and connective tissue, 20) diseases of the nervous system, 21) diseases of the respiratory system, 22) diseases of the skin and subcutaneous tissue, 23) endocrine, nutritional and metabolic diseases, 24) gingivitis, 25) hypertension, 26) injury, poisoning and certain other consequences of external causes, 27) mental and behavioral disorders, 28) neoplasms, 29 periodontitis, 30) pregnancy, childbirth and the puerperium, 31) pulmonary tuberculosis, 32) rheumatoid arthritis, 33) Sjogren’s syndrome, 34) stroke or neural conditions, 35) symptoms, signs and abnormal clinical and laboratory findings, not elsewhere classified, 36) systemic lupus erythematosus, 37) systemic sclerosis, 38) organ transplant, and 39) other unspecified factors influencing health status and contact with health services.

## Supplementary Section 4. COVID-19 severity, criticality, and fatality classification

Classification of COVID-19 case severity (acute-care hospitalizations) [27], criticality (intensive-care-unit hospitalizations) [27], and fatality [28] followed the World Health Organization (WHO) guidelines. Assessments were made by trained medical personnel independent of study investigators and using individual chart reviews, as part of a national protocol applied to every hospitalized COVID-19 patient. Following the national protocol, every individual with a SARS-CoV-2-positive test and a concurrent COVID-19 hospital admission was assessed for infection severity at regular intervals until discharge or death, throughout the pandemic, regardless of the length of hospital stay [29]. We classified individuals who progressed to severe, critical, or fatal COVID-19 between the time of the documented infection and the end of the study based on their worst outcome, starting with death [28], followed by critical disease [27], and then severe disease [27].

## Severe COVID-19

Severe COVID-19 disease was defined per WHO classification as a SARS-CoV-2 infected person with "oxygen saturation of <90% on room air, and/or respiratory rate of >30 breaths/minute in adults and children >5 years old (or ≥60 breaths/minute in children <2 months old or ≥50 breaths/minute in children 2-11 months old or ≥40 breaths/minute in children 1–5 years old), and/or signs of severe respiratory distress (accessory muscle use and inability to complete full sentences, and, in children, very severe chest wall indrawing, grunting, central cyanosis, or presence of any other general danger signs)" [27]. Detailed WHO criteria for classifying SARS-CoV-2 infection severity can be found in the WHO technical report [27].

## Critical COVID-19

Critical COVID-19 disease was defined per WHO classification as a SARS-CoV-2 infected person with "acute respiratory distress syndrome, sepsis, septic shock, or other conditions that would normally require the provision of life sustaining therapies such as mechanical ventilation (invasive or non-invasive) or vasopressor therapy" [27]. Detailed WHO criteria for classifying SARS-CoV-2 infection criticality can be found in the WHO technical report [27].

## Fatal COVID-19

COVID-19 death was defined per WHO classification as "a death resulting from a clinically compatible illness, in a probable or confirmed COVID-19 case, unless there is a clear alternative cause of death that cannot be related to COVID-19 disease (e.g. trauma). There should be no period of complete recovery from COVID-19 between illness and death. A death due to COVID- 19 may not be attributed to another disease (e.g. cancer) and should be counted independently of preexisting conditions that are suspected of triggering a severe course of COVID-19". Detailed WHO criteria for classifying COVID-19 death can be found in the WHO technical report [28].

## Supplementary Section 5. Further details on the matching process

Matching by calendar time was implemented to ensure that matched pairs were present in Qatar during the same period. Individuals in the two-dose analysis who received their second vaccine dose in a specific calendar week were matched to unvaccinated individuals who had a record of a SARS-CoV-2-negative test during the same week.

Similarly, individuals in the three-dose analysis who received their third vaccine dose in a specific calendar week were matched to individuals in the two-dose cohort who had a record of a SARS-CoV-2-negative test during the same week. Cohorts were additionally matched by calendar week of the second vaccine dose to ensure that paired individuals received their primary-series vaccination at the same time.

In the natural-infection analysis, individuals who had their primary infection in a specific calendar week were matched to uninfected individuals who had a record of a SARS-CoV-2- negative test during the same week.

In all analyses, individuals who were tested after death, had an unascertained or discrepant death date, or died before the start of follow-up were excluded.

The matching approach for this study aimed to balance observed confounders that could potentially affect the risk of infection across the exposure groups [2, 26, 30–32]. The matching factors were selected based on findings of earlier COVID-19 epidemiologic studies on Qatar’s population [2, 6, 11, 14, 21, 24, 33].

The matching algorithm was implemented using *ccmatch* command in Stata 18.0 supplemented with conditions to retain only controls that fulfilled the eligibility criteria, and was iterated using loops with as many replications as needed until exhaustion (i.e., no more matched pairs could be identified).

According to our study designs and matching approaches, individuals in the vaccine effectiveness studies may have contributed follow-up time as part of the matched control cohort before their vaccination status changed. Subsequently, based on their new vaccination status, they could have contributed follow-up time as part of the two-dose cohort or three-dose cohort. Similarly, in the natural-infection effectiveness study, individuals may have contributed follow-up time as part of the matched uninfected cohort and, upon infection, subsequently as part of the primary-infection cohort, reflecting the change in their infection status.

**Supplementary Fig. 1.**
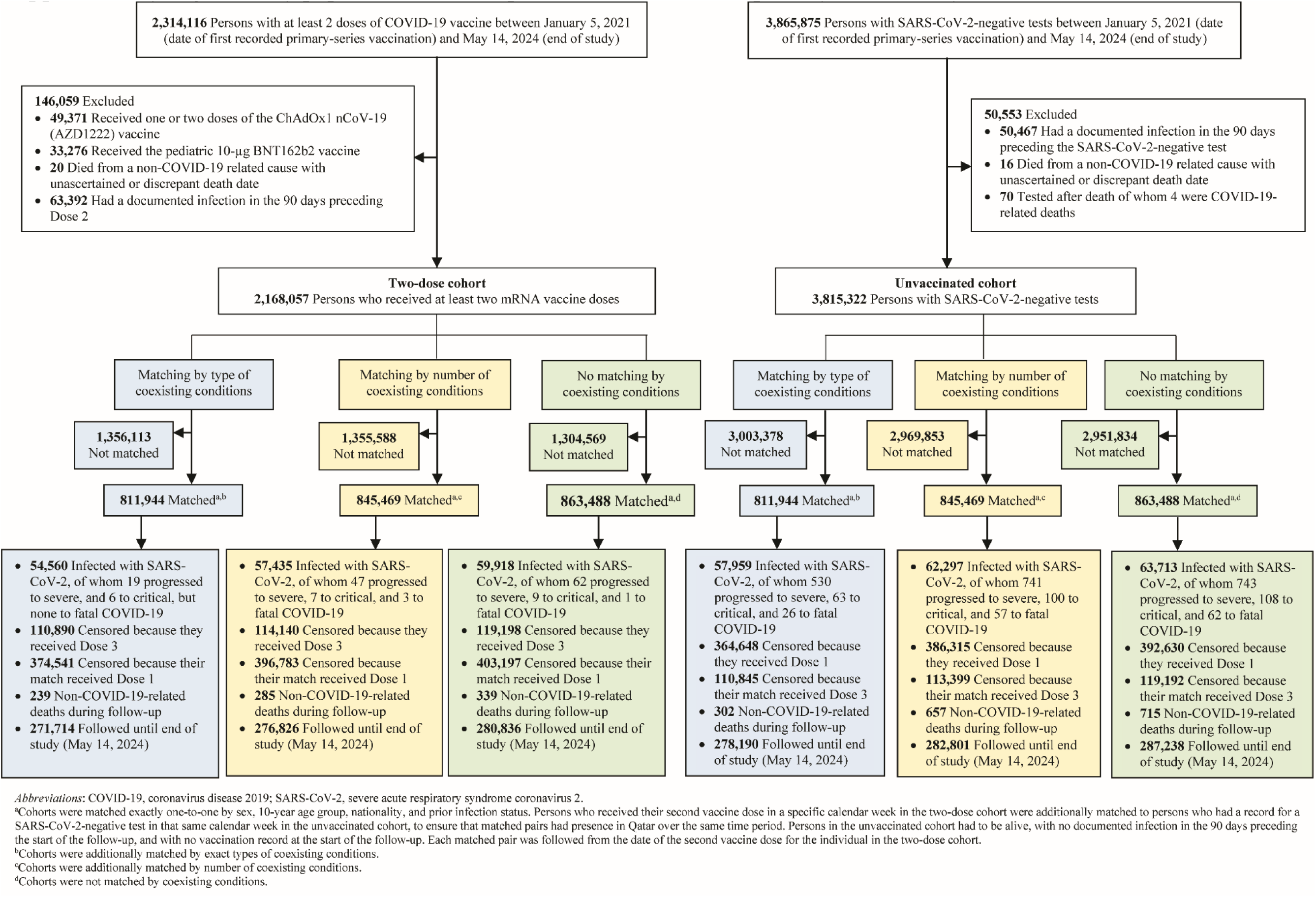
Study population selection process in the two-dose (primary-series) analysis.

**Supplementary Fig. 2.**
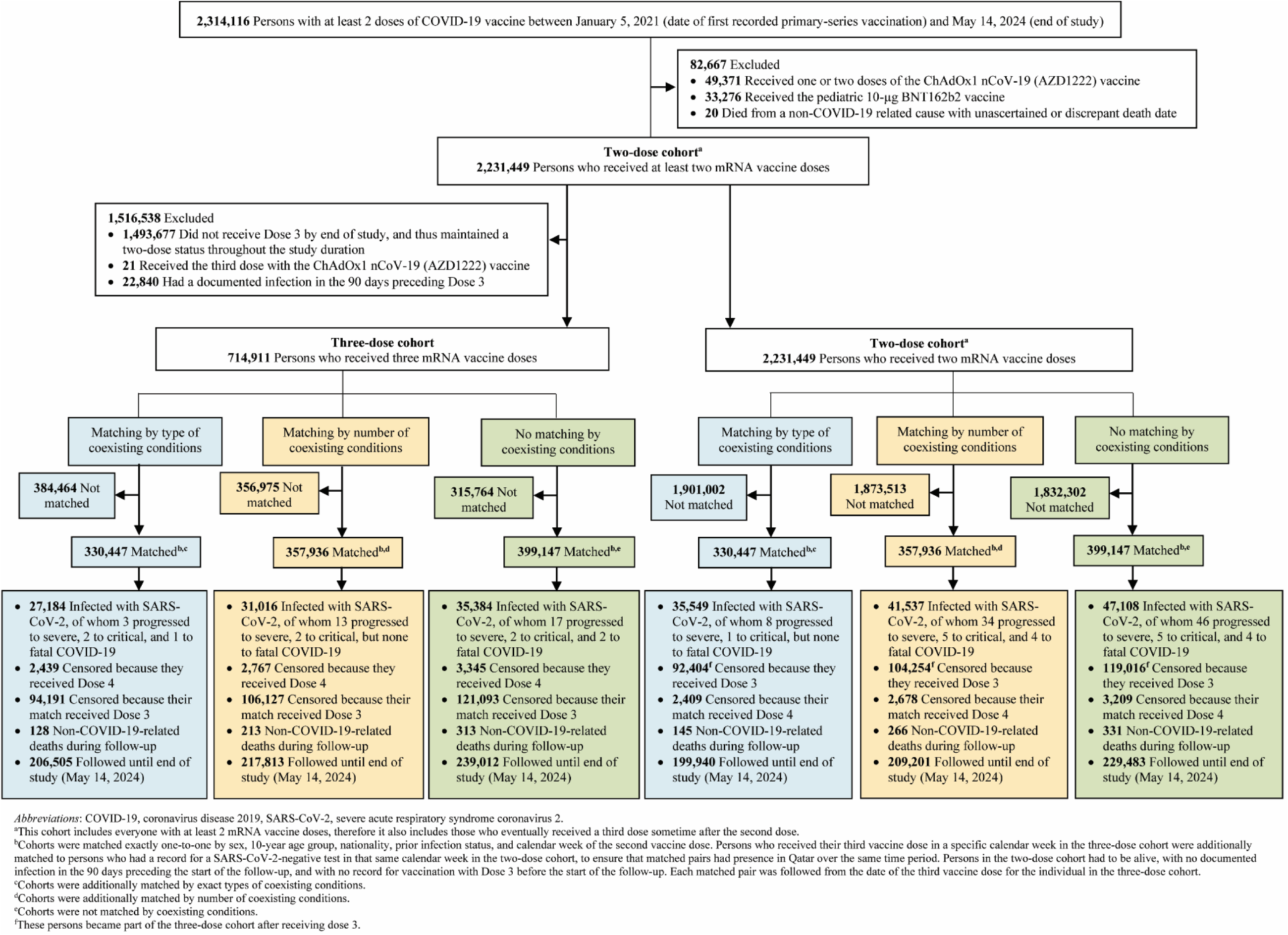
Study population selection process in the three-dose (booster) analysis.

**Supplementary Fig. 3.**
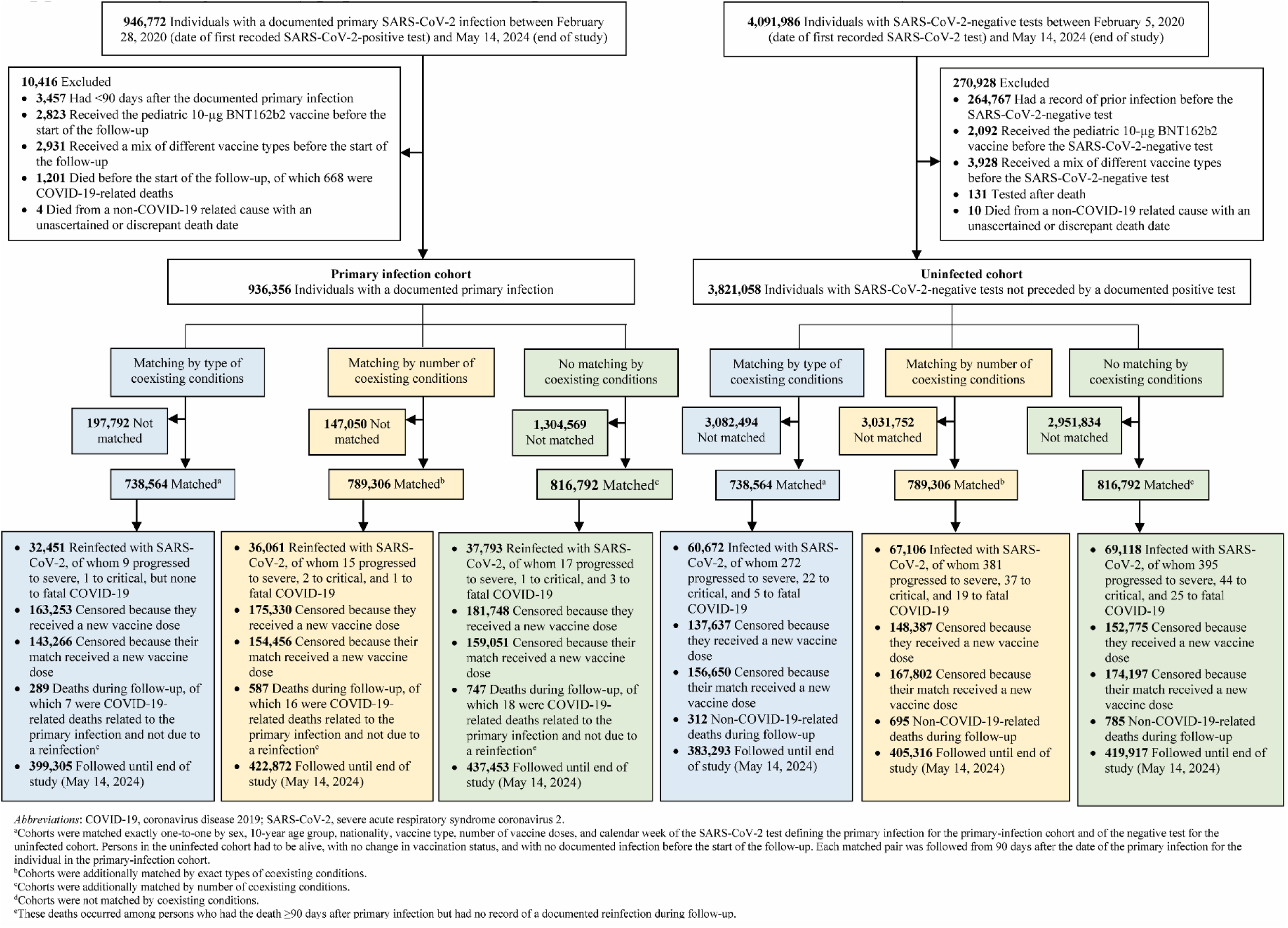
Study population selection process in the natural-infection analysis.

**Supplementary Table 1.** Strengthening the Reporting of Observational Studies in Epidemiology (STROBE) checklist for cohort studies.

|  | Item No | Recommendation | Main Text page |
| --- | --- | --- | --- |
| Title and abstract | 1 | (a) Indicate the study’s design with a commonly used term in the title or the abstract | Abstract |
|  |  | (b) Provide in the abstract an informative and balanced summary of what was done and what was found | Abstract |
| Introduction |  |  |  |
| Background/rationale | 2 | Explain the scientific background and rationale for the investigation being reported | Introduction |
| Objectives | 3 | State specific objectives, including any prespecified hypotheses | Introduction |
| Methods |  |  |  |
| Study design | 4 | Present key elements of study design early in the paper | Methods (‘Study population, data sources, and vaccination’ & ‘Study design’) |
| Setting | 5 | Describe the setting, locations, and relevant dates, including periods of recruitment, exposure, follow-up, and data collection | Methods (‘Study population, data sources, and vaccination’, ‘Study design’, & ‘Cohorts’ follow-up’) & Supplementary Figs. 1-3 in Supplementary Appendix |
| Participants | 6 | (a) Give the eligibility criteria, and the sources and methods of selection of participants. Describe methods of follow-up<br>(b) For matched studies, give matching criteria and number of exposed and unexposed | Methods (‘Inclusion criteria and matching’ & ‘Cohorts’ follow-up’), & Supplementary Figs. 1-3 & Supplementary Section 5 in Supplementary Appendix |
| Variables | 7 | Clearly define all outcomes, exposures, predictors, potential confounders, and effect modifiers. Give diagnostic criteria, if applicable | Methods (‘Study design’, ‘Inclusion criteria and matching’ & ‘Cohorts’ follow-up’, & ‘Statistical analysis’), Table 1, & Supplementary Sections 1-5 in Supplementary Appendix |
| Data sources/measurement | 8* | For each variable of interest, give sources of data and details of methods of assessment (measurement). Describe comparability of assessment methods if there is more than one group | Methods (‘Study population, data sources, and vaccination’, ‘Study design’, & ‘Statistical analysis’), Table 1, & Supplementary Sections 1-4 in Supplementary Appendix |
| Bias | 9 | Describe any efforts to address potential sources of bias | Methods (‘Inclusion criteria and matching’, ‘Cohorts’ follow-up’, & ‘Statistical analysis’) |
| Study size | 10 | Explain how the study size was arrived at | Supplementary Figs. 1-3 in Supplementary Appendix |
| Quantitative variables | 11 | Explain how quantitative variables were handled in the analyses. If applicable, describe which groupings were chosen and why | Methods (‘Inclusion criteria and matching’ & ‘Statistical analysis’) & Tables 1, 3 & 4 |
| Statistical methods | 12 | (a) Describe all statistical methods, including those used to control for confounding | Methods (‘Statistical analysis’) |
|  |  | (b) Describe any methods used to examine subgroups and interactions | Methods (‘Statistical analysis’) |
|  |  | (c) Explain how missing data were addressed | Not applicable, see Methods (‘Study population, data sources, and vaccination’) & Supplementary Section 1 in Supplementary Appendix |
|  |  | (d) If applicable, explain how loss to follow-up was addressed | Not applicable, see Methods (‘Study population, data sources, and vaccination’) & Supplementary Section 1 in Supplementary Appendix |
|  |  | (e) Describe any sensitivity analyses | Methods (‘Statistical analysis’) |
| Results |  |  |  |
| Participants | 13* | (a) Report numbers of individuals at each stage of study—eg numbers potentially eligible, examined for eligibility, confirmed eligible, included in the study, completing follow-up, and analysed<br>(b) Give reasons for non-participation at each stage<br>(c) Consider use of a flow diagram | Results (‘Two-dose analysis’, paragraph 1, ‘Three-dose analysis’, paragraph 1, & ‘Natural-infection analysis’, paragraph 1), Tables 1, 3 & 4, & Supplementary Figs. 1-3 in Supplementary Appendix |
| Descriptive data | 14 | (a) Give characteristics of study participants (eg demographic, clinical, social) and information on exposures and potential confounders | Tables 1, 3 & 4 |
|  |  | (b) Indicate number of participants with missing data for each variable of interest | Not applicable, see Methods ('Study population, data sources, and vaccination') & Supplementary Section 1 in Supplementary Appendix |
|  |  | (c) Summarise follow-up time (eg, average and total amount) | Tables 2 & 5 |
| Outcome data | 15 | Report numbers of outcome events or summary measures over time | Tables 2 & 5, & Supplementary Figs. 1-3 in Supplementary Appendix |
| Main results | 16 | (a) Give unadjusted estimates and, if applicable, confounder-adjusted estimates and their precision (eg, 95% confidence interval). Make clear which confounders were adjusted for and why they were included | Results ('Two-dose analysis', paragraphs 2-3 & 5, 'Three-dose analysis', paragraphs 2-3 & 5, & 'Natural-infection analysis', paragraphs 2-3 & 5), Fig. 1, & Tables 2 & 5 |
|  |  | (b) Report category boundaries when continuous variables were categorized | Tables 1, 3 & 4 |
|  |  | (c) If relevant, consider translating estimates of relative risk into absolute risk for a meaningful time period | Not applicable |
| Other analyses | 17 | Report other analyses done—eg analyses of subgroups and interactions, and sensitivity analyses | Results ('Two-dose analysis', paragraph 4, 'Three-dose analysis', paragraph 4, & 'Natural-infection analysis', paragraph 4), Fig. 1, & Tables 2 & 5 |
| Discussion |  |  |  |
| Key results | 18 | Summarise key results with reference to study objectives | Discussion, paragraphs 1-6 |
| Limitations | 19 | Discuss limitations of the study, taking into account sources of potential bias or imprecision. Discuss both direction and magnitude of any potential bias | Discussion, paragraphs 7-9 |
| Interpretation | 20 | Give a cautious overall interpretation of results considering objectives, limitations, multiplicity of analyses, results from similar studies, and other relevant evidence | Discussion, paragraph 11 |
| Generalisability | 21 | Discuss the generalisability (external validity) of the study results | Discussion, paragraphs 7-9 |
| Other information |  |  |  |
| Funding | 22 | Give the source of funding and the role of the funders for the present study and, if applicable, for the original study on which the present article is based | Funding |

**Supplementary Table 2.** Sensitivity analysis. Hazard ratios for incidence of severe, critical, or fatal COVID-19 and of non-COVID- 19 death among individuals who are clinically vulnerable to severe COVID-19 in the A) two-dose (primary-series) analysis, B) three-dose (booster) analysis, and C) natural-infection analysis using different criteria for matching by coexisting conditions.

| Sensitivity analysis | Matching by coexisting condition types |  | Matching by number of coexisting conditions |  | No matching by coexisting conditions |  |
| --- | --- | --- | --- | --- | --- | --- |
| A) Two-dose analysis | Two-dose<sup>a,b</sup> | Unvaccinated<sup>a,b</sup> | Two-dose<sup>a,c</sup> | Unvaccinated<sup>a,c</sup> | Two-dose<sup>a,d</sup> | Unvaccinated<sup>a,d</sup> |
| *Severe, critical, or fatal COVID-19* |  |  |  |  |  |  |
| Adjusted hazard ratio (95% CI)<sup>e</sup> | 0.03 (0.02-0.06) |  | 0.08 (0.06-0.10) |  | 0.08 (0.06-0.10) |  |
| Vaccine effectiveness (95% CI) | 96.7 (94.0-98.2) |  | 92.4 (89.7-94.4) |  | 92.2 (89.5-94.2) |  |
| *Non-COVID-19 death* |  |  |  |  |  |  |
| Adjusted hazard ratio (95% CI)<sup>f</sup> | 0.54 (0.41-0.71) |  | 0.22 (0.18-0.27) |  | 0.24 (0.20-0.28) |  |
| B) Three-dose analysis | Three-dose<sup>g,b</sup> | Two-dose<sup>g,b</sup> | Three-dose<sup>g,c</sup> | Two-dose<sup>g,c</sup> | Three-dose<sup>g,d</sup> | Two-dose<sup>g,d</sup> |
| *Severe, critical, or fatal COVID-19* |  |  |  |  |  |  |
| Adjusted hazard ratio (95% CI)<sup>h</sup> | 0.16 (0.02-1.33) |  | 0.30 (0.14-0.60) |  | 0.30 (0.16-0.54) |  |
| Vaccine effectiveness (95% CI) | 84.1 (-25.0-98.1) |  | 70.5 (39.7-85.6) |  | 70.3 (45.5-83.8) |  |
| *Non-COVID-19 death* |  |  |  |  |  |  |
| Adjusted hazard ratio (95% CI)<sup>i</sup> | 0.71 (0.48-1.04) |  | 0.61 (0.49-0.77) |  | 0.71 (0.59-0.87) |  |
| C) Natural-infection analysis | Primary infection<sup>j,b</sup> | Uninfected<sup>j,b</sup> | Primary infection<sup>j,c</sup> | Uninfected<sup>j,c</sup> | Primary infection<sup>j,d</sup> | Uninfected<sup>j,d</sup> |
| *Severe, critical, or fatal COVID-19* |  |  |  |  |  |  |
| Adjusted hazard ratio (95% CI)<sup>k</sup> | 0.04 (0.02-0.11) |  | 0.05 (0.03-0.10) |  | 0.05 (0.03-0.09) |  |
| Natural-infection effectiveness (95% CI) | 95.5 (89.1-98.2) |  | 94.7 (90.5-97.0) |  | 94.7 (90.6-97.1) |  |
| *Non-COVID-19 death* |  |  |  |  |  |  |
| Adjusted hazard ratio (95% CI)<sup>l</sup> | 0.78 (0.60-1.00) |  | 0.68 (0.60-0.78) |  | 0.76 (0.67-0.86) |  |

Abbreviations: CI, confidence interval; COVID-19, coronavirus disease 2019; SARS-CoV-2, severe acute respiratory syndrome coronavirus 2.
<sup>a</sup>Cohorts were matched exactly one-to-one by sex, 10-year age group, nationality, and prior infection status. Persons who received their second vaccine dose in a specific calendar week in the two-dose cohort were additionally matched to persons who had a record for a SARS-CoV-2-negative test in that same calendar week in the unvaccinated cohort, to ensure that matched pairs had presence in Qatar over the same time period. Persons in the unvaccinated cohort had to be alive, with no documented infection in the 90 days preceding the start of the follow-up, and with no vaccination record at the start of the follow-up.
<sup>b</sup>Cohorts were additionally matched by exact types of coexisting conditions.
<sup>c</sup>Cohorts were additionally matched by number of coexisting conditions.
<sup>d</sup>Cohorts were not matched by coexisting conditions.
<sup>e</sup>Adjusted for sex, 10-year age group, nationality, number of coexisting conditions, prior infection status, calendar week of the second vaccine dose for the two-dose cohort or SARS-CoV-2-negative test for the unvaccinated cohort, and testing rate.
<sup>f</sup>Adjusted for sex, 10-year age group, nationality, number of coexisting conditions, prior infection status, and calendar week of the second vaccine dose for the two-dose cohort or SARS-CoV-2-negative test for the unvaccinated cohort.
<sup>g</sup>Cohorts were matched exactly one-to-one by sex, 10-year age group, nationality, prior infection status, and calendar week of the second vaccine dose. Persons who received their third vaccine dose in a specific calendar week in the three-dose cohort were additionally matched to persons who had a record for a SARS-CoV-2-negative test in that same calendar week in the two-dose cohort, to ensure that matched pairs had presence in Qatar over the same time period. Persons in the two-dose cohort had to be alive, with no documented infection in the 90 days preceding the start of the follow-up, and with no record for vaccination with Dose 3 before the start of the follow-up.
<sup>h</sup>Adjusted for sex, 10-year age group, nationality, number of coexisting conditions, prior infection status, calendar week of the second vaccine dose, and testing rate.
<sup>i</sup>Adjusted for sex, 10-year age group, nationality, number of coexisting conditions, prior infection status, and calendar week of the second vaccine dose.

